# SEIHCRD Model for COVID-19 spread scenarios, disease predictions and estimates the basic reproduction number, case fatality rate, hospital, and ICU beds requirement

**DOI:** 10.1101/2020.07.24.20161752

**Authors:** Avaneesh Singh, Manish Kumar Bajpai

## Abstract

We have proposed a new mathematical method, SEIHCRD-Model that is an extension of the SEIR-Model adding hospitalized and critical two-compartments. SEIHCRD model has seven compartments: susceptible (S), exposed (E), infected (I), hospitalized (H), critical (C), recovered (R), and deceased or death (D), collectively termed SEIHCRD. We have studied COVID-19 cases of six countries, where the impact of this disease in the highest are Brazil, India, Italy, Spain, the United Kingdom, and the United States. SEIHCRD model is estimating COVID-19 spread and forecasting under uncertainties, constrained by various observed data in the present manuscript. We have first collected the data for a specific period, then fit the model for death cases, got the values of some parameters from it, and then estimate the basic reproduction number over time, which is nearly equal to real data, infection rate, and recovery rate of COVID-19. We also compute the case fatality rate over time of COVID-19 most affected countries. SEIHCRD model computes two types of Case fatality rate one is CFR daily and the second one is total CFR. We analyze the spread and endpoint of COVID-19 based on these estimates. SEIHCRD model is time-dependent hence we estimate the date and magnitude of peaks of corresponding to the number of exposed cases, infected cases, hospitalized cases, critical cases, and the number of deceased cases of COVID-19 over time. SEIHCRD model has incorporated the social distancing parameter, different age groups analysis, number of ICU beds, number of hospital beds, and estimation of how much hospital beds and ICU beds are required in near future.

## 1 Introduction

The world is fighting against a new enemy in these days, which is the COVID-19 virus. The COVID-19 disease, known as coronavirus disease in 2019. The virus has spread quickly in the world since its first appearance in China. We all are fighting every day against all the economical and social implications because of this virus and most of the countries are facing this new enemy in the Western countries.

China has reported a pneumonia outbreak in Wuhan in December 2019 [1]. This outbreak was linked to a novel strain of coronavirus on 31 December 2019 [2], which has given the tentative name 2019-nCoV by the World Health Organization (WHO) [3-5], later renamed SARS-CoV-2 by the International Committee on Taxonomy of Viruses. Tyrell and Bynoe [6] first describe Coronavirus in 1966. The World Health Organization announced these outbreaks as a public health emergency of international on 30 January and a pandemic on 11 March 2020 [7, 8].

This pandemic has reported more than 5.59 million cases of COVID-19 have been reported in more than 188 countries and territories and more than 2.28 million people have recovered from the virus and more than 350,000 deaths as of 27 May 2020 [9]. World Health Organization has not yet given official validity to any of the vaccines for COVID-19 [10]. United States has permitted to use antiviral remdesivir for severe patients of COVID-19 [11]. COVID-19 has transmitted among people through close contact, small droplets via coughing, talking, and sneezing [12-14]. The droplets usually not stay in the air more time hence it falls on to the ground instead of traveling in the air [12]. People have also be infected by contacting the infected substance and then contacting their faces with unwashed hands [12, 13].

COVID-19, the most common symptoms are fever, it includes cough, fatigue, shortness of breath, and loss of sense of smell [12, 15], and in some cases, pneumonia and acute respiratory distress syndrome symptoms find in COVID-19 [16]. Lungs are the most affected part of the body by COVID-19 [17]. Around one out of every six people who get COVID-19 becomes seriously ill and develops difficulty breathing [18]. World Health Organization suggested some preventive measures that include hand washing maintain distance from other people (social distancing), covering one’s mouth while coughing, always use a face mask in a public place, and follow self-isolation when anyone feels infected [12,19]. Many countries follow lockdown, restrictions on travel, work from home, and facility closure control to spread the virus. We have to follow staying at home, avoid public places, keep the distance from others, sanitize your hand regularly and wash your hand for at least 20 seconds, avoid touching face, eyes, hairs, nose, and mouth with unwashed hands control to spread this virus [20-22].

COVID-19 has caused the global recession and it affects social life [23]. COVID-19 has affected many sporting activities; financial, religious, and political activities have been rescheduled or canceled [23, 25]. This tragedy also has some good consequences, the emission of pollutants and greenhouse gases are reduced [26, 27]. China has reported around 80 percent of deaths in those over 60 years of age and 75 percent in chronic health conditions, including heart disease and diabetes, as of 5 February [28]. Wuhan is the city where the first death case has reported on 9 January 2020 [29]. The Philippines is the country where the first confirmed case death outside China has reported on 1 February [30], and the first confirmed case reported outside of Asia on 14 February in France [31]. The WHO and Chinese officials have reported human-to-human transmission by 20 January 2020[32, 33]. Johns Hopkins University estimates that from the global death to the case ratio is 6.3 percent (350,458 fatalities per 5,591,067 cases) as of 27 May 2020 [9].

Many smartphone applications have been developed for voluntary use. These application uses Bluetooth to log a user’s proximity to other smartphones. Users get a warning message when they have been a direct touch with someone who has recently tested positive for COVID-19[34]. Italy has reported its first confirmed case on 31 January 2020, two visitors come from China [35]. WHO declared Europe as the active hub of a pandemic on 13 March 2020[36]. Italy has surpassed China with the most number of fatalities on 19 March 2020 [37]. The United States become the most number of cases reported in the world by 26 March [38]. European travelers are the main source of spreaders in New York [39]. Nearly 3.9 billion people had under some form of lockdown by the first week of April [40].

COVID-19 has a profound impact on the economy of all countries. Global stock markets fell on 24 February 2020 because of the rapid increase in the number of cases of COVID-19 across mainland China [41, 42]. The world has seen the financial crisis first-time a huge decline in stock markets after 2008 and it crashed in March 2020 [43-45] and because of this pandemic, many financial deals are being canceled or postponed [46].

This paper proposed a new mathematical model to study the dynamics of transmission and control of the COVID-19 pandemic in most affected countries. The SEIR model is a widely used epidemiological model based on the SIR model, which was given, by Kermack and Mckendrick [47]. Kermack and Mckendrick’s epidemic model has been very effective in forecasting outbreak behavior quite close to that seen in several reported epidemics [48]. After Kermack and Mckendrick’s manuscript, many epidemic mathematical models have been developed stochastic models, discrete-time models, continuous-time models, and diffusion models. Compartmental models primarily depend on differential equations. These models are normally subdivided into many compartments (i.e., Susceptible, Infected, Recovered, Critical, Death, etc.). Compartmental models focused on basic law defining the transition of people from one compartment to another. These Models use mathematics to find the different parameters to estimate the effect of different interventions. Modeling can help determine which intervention to stop and which to continue and can predict the future [49]. The model can predict how disease spreads, what will be the lifetime of any epidemic, how much people infected, recovered, and deceased. It can also estimate many parameters like reproduction numbers etc. These models are used in many applications nowadays mostly in epidemiology, and many other fields like economics, politics, social science, and in the medical field. Epidemiological models such as SEIR provide a valuable computational approach to understanding the microscopic view of the spread of disease. These models play an important role to measure possible strategies for the control and mitigation of infectious diseases [49-51]. These models are useful in cases where disease dynamics are not unclear. It estimates the number of cases in worst and best-case scenarios. Mathematical models are also helpful in understanding the scenario for spreading the disease[52-54].

The SEIR model failed to estimate the spread where preventive measures are adopted such as social distancing, different age groups, number of ICU beds, number of hospital beds, and mortality rate. The mathematical model is very helpful to predict the disease outbreak of COVID-19 and the SIR model and SEIR model have been widely used for prediction. SEIHCRD model is the modification of the SEIR model. The present manuscript encompasses a new modified SEIR model which estimates the spread of COVID-19 spread, including all the parameters mentioned above.

We have proposed a model for forecasting under uncertainties, constrained by various observed data. The model allows studying scenarios for the progress of disease spreading. SEIHCRD model estimates the date and magnitude of peaks of corresponding to exposed people, the number of infected people, the number of people hospitalized, the number of people admitted in ICUs, and the number of death of COVID-19.

The proposed model has incorporated the social distancing parameter, different age groups analysis, number of ICU beds, number of hospital beds and lockdown effect gives a more accurate estimation of how much hospital beds and intensive care unit (ICU) beds are required in near future. Also, the proposed model analyzes the reproduction number of most affected countries and when the disease goes settle. The basic reproduction number for COVID-19 in January was between 1.4 and 2.5 [55], but later analysis showed that this can vary from 3.8 to 8.9 [56]. As of last May 2020, the spread of disease in most of the countries is either stable or decreasing [57].

The structure of this paper is as follows; Section 1 introduces COVID-19 and explains the significance of this research. The SEIHCRD model for COVID-19 spread estimation; predictive modeling has been presented in section 2. Section 3 has discussed the data analysis and parameter estimation used for validating proposed mathematical models. Results and discussions of the proposed model have been discussed in section 4. Finally, the conclusion and some future works have been discussed in Section 5.

## 2 Proposed methodology

The SIR and SEIR compartmental models are the most commonly used mathematical method for infectious disease. SEIHCRD-Model is a new mathematical method that is an extension of the SEIR-Model adding death, hospitalized, and critical compartments. The proposed method and some basic compartmental models have been described below.

### 2.1 SIR Model

Mathematical modeling of infectious disease specially Coronavirus disease (COVID-19) has been a trending topic for many epidemiologists and data scientists in the last some weeks. Coronavirus disease (COVID-19) is an infectious disease hence it can spread from one member of the population to another member of the population. Kermack-McKendrick [47] has proposed the SIR (susceptible, infected, recovered) model in 1927. SIR model is separated into three population-wise compartments of Susceptible (S), Infectious (I), and Recovered (R). SIR model describes the transition of people from Susceptible to Infected and then Infected to Recovered based on underlying parameters, which controls disease dynamics in the population. Deceased cases come under the Recovered compartment in the SIR model. The SIR model is shown in Fig. 1.

**Figure 1:**
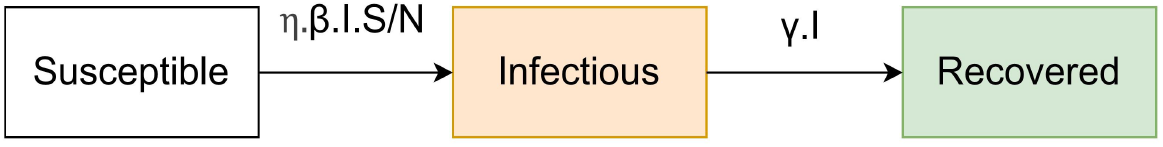
SIR Model

#### Differential Equations for Traditional SIR Model

We write the model equations that are independent of the population of the country by considering the fraction of the people in each category. The rates of transfer from one category to another are the model parameters and a set of differential equations are formed. The SIR model has been described below using the coupled ordinary differential equations:

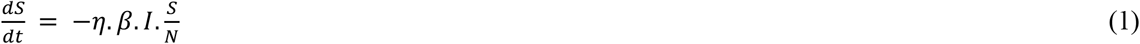

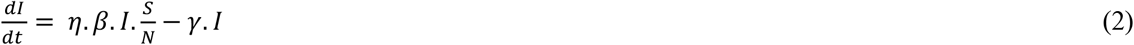

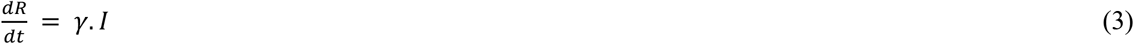

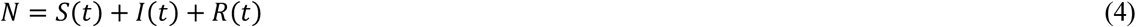

Where,

N – Total population

S – A proportion of the entire population that is healthy and have never been infected

I - A proportion of the entire population that is infected by the virus

R - A proportion of the entire population that has recovered from the infection

Where initial conditions (S (0), I (0), R (0)) are not exactly known, and these systems of ordinary differential equations are *extremely sensitive to the initial parameters. Description of some parameters are given below:*

η – social distancing factor

β – the rate of transmission of infection from susceptible to infected

γ- the rate of recovery

R_0_ – basic reproduction number

X – number of days an infected person has and can spread the diseases 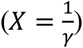

Basic reproduction number *R*_0_ *= β.X*

Therefore, from above we can write it as 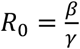

### 2.2 SEIR Model

Kermack and McKendrick [47], Beretta, and Takeuchi [58] have further developed their theories and proposed a time delay SIR model. Cooke and Driessche [59] investigated the incubation period in the spread of infectious diseases, introduced the “Exposed, *E*,“ compartment, and proposed a time delay model. SEIR model is separated into four population-wise compartments of Susceptible (S), Exposed (E), Infectious (I), and Recovered (R). SEIR model describes the transition of people from Susceptible to Exposed, then Exposed to Infected and then Infected to Recovered based on underlying parameters, which controls disease dynamics in the population. The SEIR model is shown in Fig. 2.

**Figure 2:**
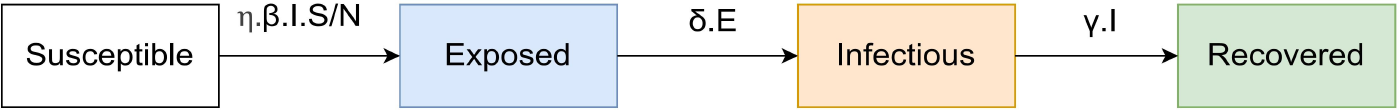
SEIR Model

Deceased cases come under the recovered compartment in the SEIR model. Deceased cases play an important role in infectious modeling hence; we have to make it a separate compartment. The extension of the SEIR model is the SEIRD model in which the Dead compartment is added. SEIR model is separated into four population-wise compartments of Susceptible (S), Exposed (E), Infectious (I), Recovered (R), and Dead (D). SEIRD model describes the transition of people from Susceptible to Exposed, then Exposed to Infected, then Infected to Recovered and Infected to Dead. The SEIRD model is shown in Fig. 3.

**Figure 3:**
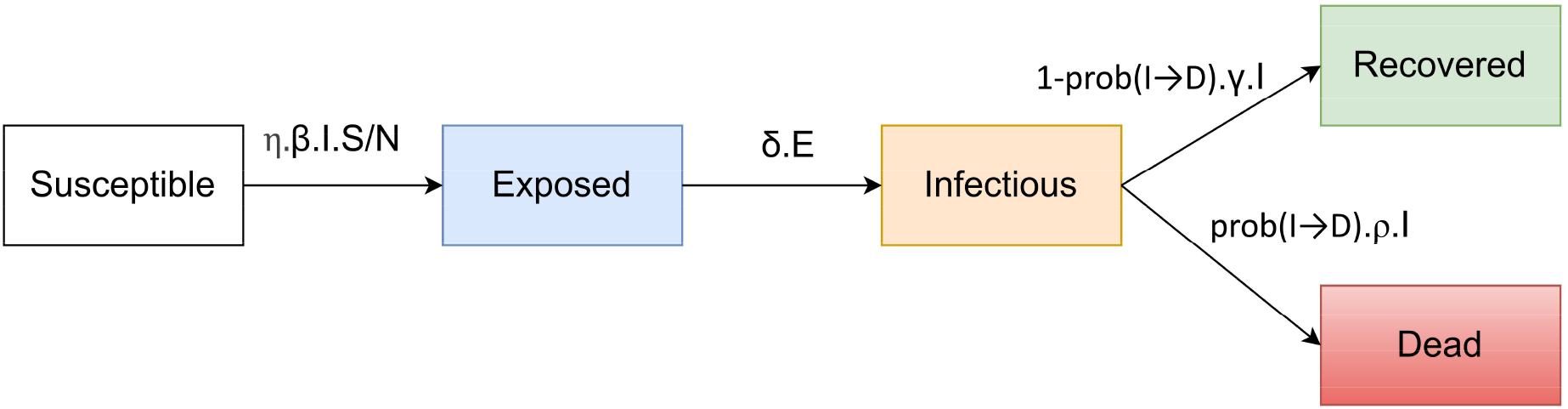
SEIRD Model

#### Differential Equations for SEIR Model and SEIRD Model

The SEIR model is described below using the coupled ordinary differential equations:

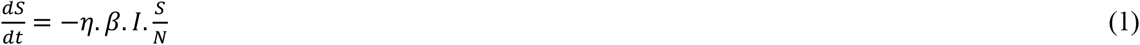

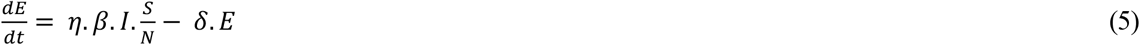

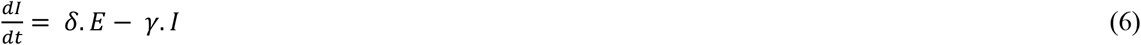

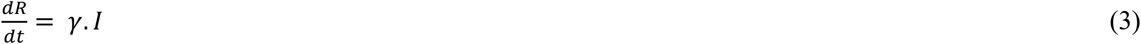

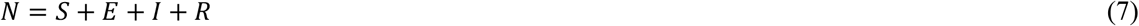

Where

E -A proportion of the entire population that is exposed to infection, transmit the infection and turn into either Symptomatic or purely Asymptomatic, and not detected

*Description of some parameters used in the SEIR model is given below:*

*β-*the rate of transmission of infection from susceptible to exposed

*δ-*the rate of transmission of infection from exposed to infectious and the rest of the parameters are the same as the SIR model.

And after adding dead compartment ordinary differential equations are change accordingly, hence coupled ordinary differential equations of SEIRD Model are given below:

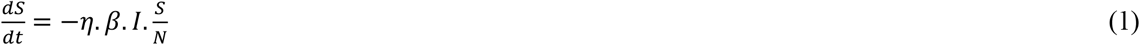

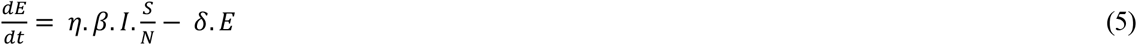

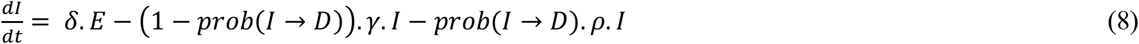

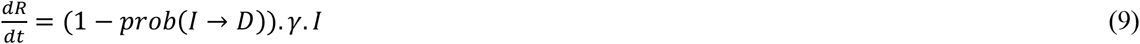

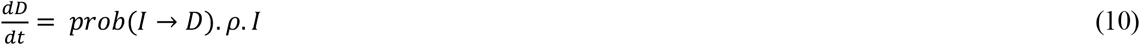

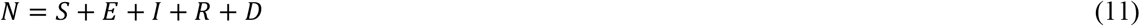

Here

D -A proportion of the entire population that is dead because of the infection.

Description of some parameters used in the SEIRD model is given below:

ρ -Median time from Infected to Death

γ -recovery period and the rest of the parameters are the same as the SIR model.

#### 2.3 SEIHCRD Model

The present manuscript encompasses two new compartments added for more accurate analysis is the Hospitalized compartment and the Critical compartment. This model allows overflowing hospitals. SEIHCRD model is separated into seven population-wise compartments of Susceptible (S), Exposed (E), Infectious (I), Hospitalized (H), Critical (C), Recovered (R), and Dead (D). SEIRD model describes the transition of people from Susceptible to Exposed, then Exposed to Infected, and then from Infected they can either Hospitalized or Recovered compartment. Of course, only infected individuals can enter the hospitalized compartment and critical compartment. From hospitalized, they can either Critical or Recover and from the Critical compartment, they can either die or recover. The SEIHCRD model is shown in Fig. 4.

**Figure 4:**
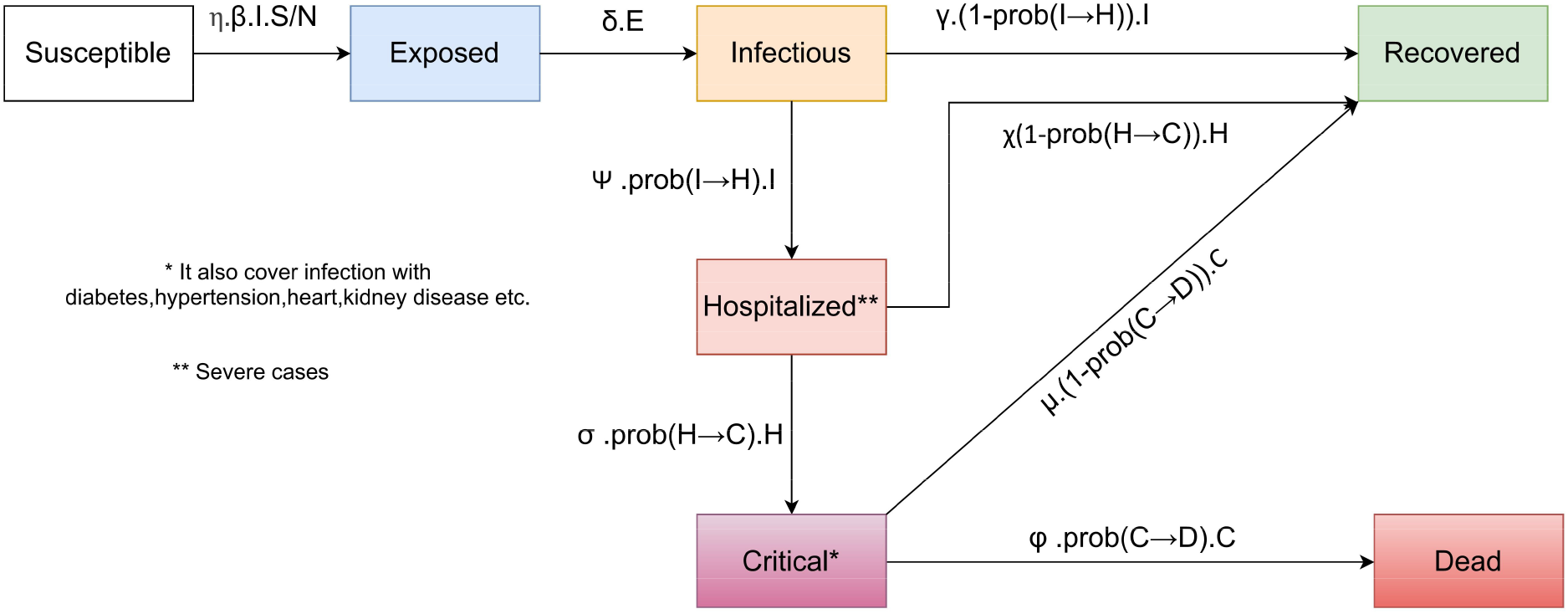
SEIHCRD Model

**Some infections are mild** like fever, cough, and may even have mild pneumonia but do not require hospitalization. These individuals may recover or in the future, they go to the hospitalized compartment.

People with a serious infection, suffer from severe pneumonia, and need hospitalization. These individuals may either recover or progress to the critical compartment. People with critical infection experience multi-organ failure and multiple disorders require treatment in an ICU. These people either recover from the disease or die from it.

*Differential Equations for* SEIHCRD *Model*

The SEIHCRD model is described below using the coupled ordinary differential equations:

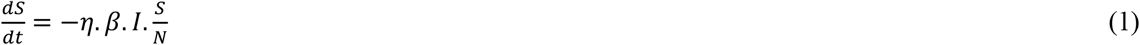

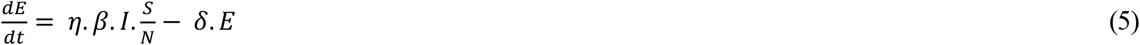

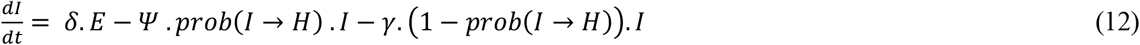

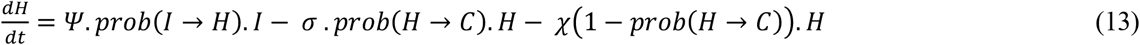

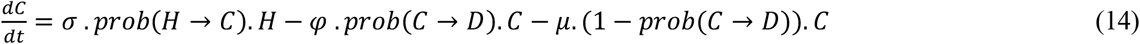

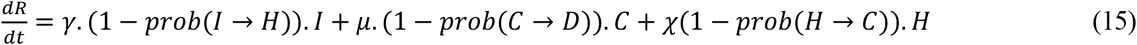

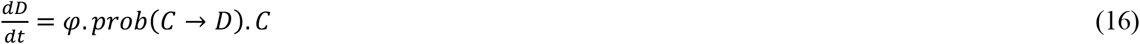

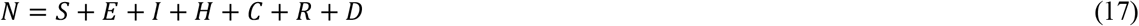

Here

H - A proportion of the entire population that is found positive in the test and hospitalized

C - A proportion of the entire population those are seriously ill and who need ICU

D - A proportion of the entire population that is dead because of the infection.

Also, the description of some parameters used in the SEIR model is given below:

*Ψ* –Median time to development of pneumonia and other symptoms for hospitalization

σ - Median time from hospital to ICU admission

φ - Median intensive care units (ICUs) length of stay

χ - Median hospital stay

μ - the recovery time of critical conditioned people

We know that every country has limited resourced of hospital beds and ICUs. Sometimes the number of critical cases is more than the number of ICUs in this condition doctors have to choose who gets treated with the limited number of resources. This manuscript accepts all these conditions as follows.

If there is a B number of ICUs and C number of critical cases then following condition occurs:

1. If the number of ICUs is more than critical cases then all patients get treated.
2. But if the number of ICUs are less than the number of critical cases (C>B) then B number of patient treated and rest (C-B) die because of shortage.

The SEIHCRD Model with critical case analysis is shown in Fig. 5.

**Figure 5:**
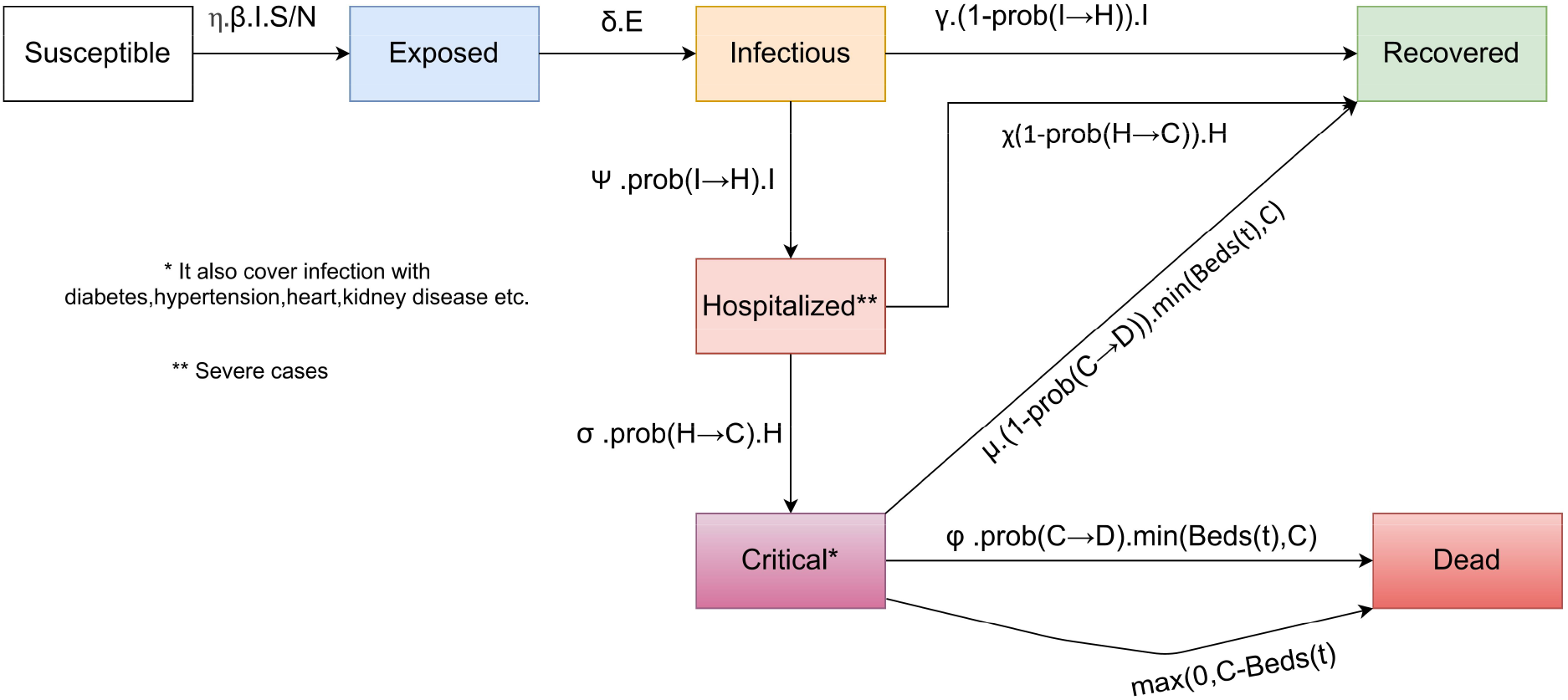
SEIHCRD Model with critical case analysis

The SEIHCRD with critical case ordinary differential equations are:

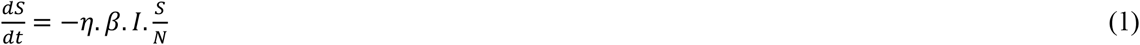

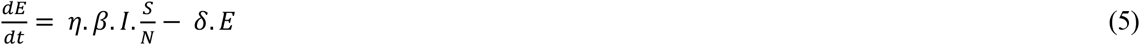

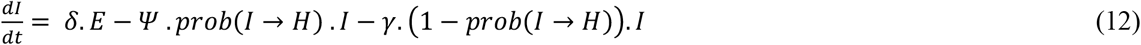

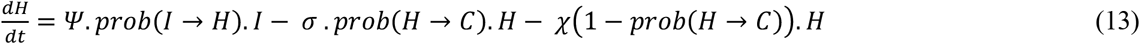

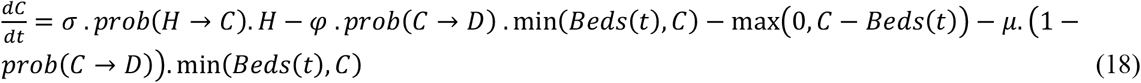

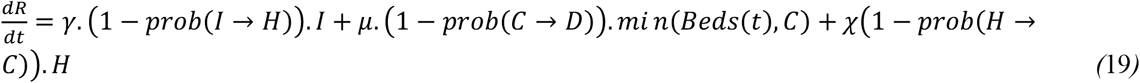

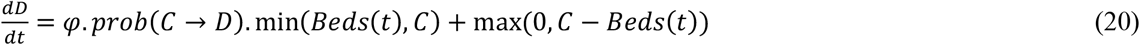

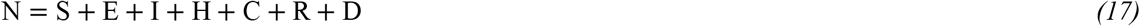

## 3 Data analysis and Parameter estimation

Compartmental models are ordinary differential equations. These ordinary differential equations have required some initial conditions and parameters to solve and characterize ODE. The prevalence of COVID-19 in the compartment, the population has influenced by the complexities of many factors. Preliminary estimation of parameters helps solve important parameters such as fatality rate and basic reproduction rate, which will help us to have a more accurate understanding of the transmission trend of COVID-19. In the present manuscript, we first collect data for a specific period, then estimate the basic reproduction number, infection rate, and recovery rate of COVID-19, and based on these estimates, we analyze the spread and endpoint of COVID-19.

η is the social distancing parameter. The social distancing parameter includes avoiding social gatherings and physical contact to avoid the spread of infectious disease. Its value lies in between zero to one. Everyone is quarantined if a lockdown is implemented and then its value is zero and they follow the normal routine without restriction then its value is one.

R (0) and R_0_ are different quantities, R (0) describes the number of recovered at *t* = 0 whereas R_0_ is the basic reproduction number that is the ratio between the rate of contacts to the rate of recovery. The basic reproduction number is the expected number, which is spread by infected people in a population where all peoples are susceptible to infected.

### R_0_ change with time

R_0_ value is change with time, whenever any country implements lockdown the value of R_0_ decreases, and when that country removes lockdown its value starts increasing again. R_0_ value has decreased when any country puts strict lockdown and after some time it goes below one. R_0_ is the ratio of β and γ in the compartmental models, hence we can say that *β = R*_0_.γ. The value of R_0_ is R_0___start_ before the lockdown and if the lockdown is imposed on day L then the value of R_0_ is decreased up to R_0___end_ and the value of β is change accordingly.

Basic reproduction number R_0_ is not constant in real life it changes with time. The basic reproduction number value has changed accordingly if social distancing imposed. The model shows the initial impact of social distancing on the reproduction number. This model use, the logistic function on it because it changes very slowly initially, its speed increase when lockdown imposed, and then it slows down in the end. The reproduction number is

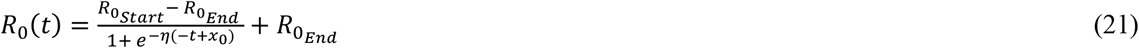

Description of parameters are given below:

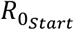 - Value of R_0_ on the first day

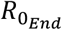 - Value of R_0_ on last day

x_0_ is the value of inflection point (i.e. the day when the value of R_0_ decline drastically)

η is the social distancing parameter.

According to the value of the social distancing parameter, it changes drastically.

### Age-dependent fatality rate

The fatality rate is not stable it depends on many things. The fatality rate affects most by one of the common reasons is the Age factor, some fatal disease, and the number of ICU beds available, etc. The fatality rate is more when the most number of people are infected, and the fatality rate and is less when infected people are less. The base fatality rate is there when fewer people are infected.

Infected people become very much sometimes, at that time we need more medical equipment for treatment, sometimes everyone is not treated due to lack of medical facility. We should know what proportion of people are currently infected, hence we describe fatality rate φ as

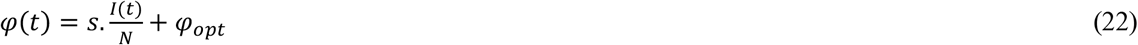

Description of parameters are given below:

s - It is arbitrary but the fixed value that controls the influence of infection (it chooses freely once and then stay constant over time)

φ_opt_ - it is the optimal fatality rate

I (t) is infected people at time t and N is the total population. The value of s controls the above equation. Therefore, carefully give the value of s.

Analysis of case fatality depends on Age group is complex. For this we separate different age groups (e.g. people aged 0 to 9, aged 10-19,…..aged 90-100). for age group analysis we need two things.

1. Fatality rates by age group,
2. The proportion of the total population is in that age group.

The fatality rate is high if the population of old people is more, and if the proportion of young people is more, then the fatality rate is low. The fatality rate has calculated by:

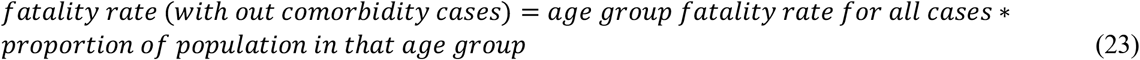

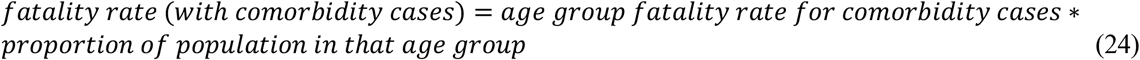

So the overall fatality rate is a combination of the above two cases then,

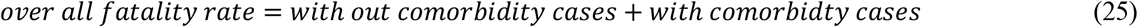

In the present manuscript, all the comorbidity cases are considered as critical cases. The fatality rate for all cases is shown in Tab. 3, and the proportion of age groups is shown in Fig. 14.

### Hospitalized and critical cases analysis

Every country has limited resourced of hospital beds and ICUs. Sometimes the number of critical cases are more than the number of ICUs in this condition doctors have to choose who gets treated with a limited number of resources. This manuscript accepts all these conditions as follows. If there is a B number of ICUs and C number of critical cases then following condition occurs:

1. People with a serious infection, suffer from severe pneumonia and need hospitalization and hospital bed are available then they treated in the hospital but sometimes beds are not available due to shortage then they have to wait till the hospital bed is empty or self-quarantine in their home.
2. If the number of ICUs is more than critical cases then all patients are treated.
3. But if the number of ICUs are less than the number of critical cases (C>B) then B number of patient treated and rest (C-B) die because of shortage

The premise for hospital beds and ICUs is that countries respond and start constructing clinics and opening the rooms, etc., while the virus spreads. Hence, over time, the number of hospital beds and ICUs is increasing. We can model the number of beds as:

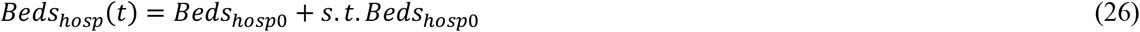

Description of parameters are given below:

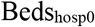 - the total amount of Hospital beds available

s - some scaling factor,

and for ICUs

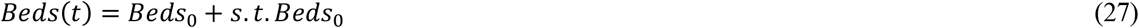

Description of parameters are given below:

Beds_0_ - the total amount of ICU beds available

s - some scaling factor

As shown in the formula number of beds increases s times per day hence the value of s, plays an important role to calculate how many numbers of beds increase daily according to this patient’s serve in the hospitals.

### Fitting the model to find some important parameters value

In this manuscript, we are focus on fitting the SEIHCRD Model with time-dependent basic reproduction number and age group based fatality rate, Hospital beds and ICUs with real COVID-19 data, that come close to the real data to find the parameters for our model that produce the possible prediction which is helpful in the future development.

All the experimental studies have been performed on the PYTHON platform using some important libraries i.e. pandas, NumPy, lmfit, ode, etc. The machine used for performing simulation work has Processor Intel(R) Core(TM) i7-5500U CPU @ 2.40 GHz, 2401 MHz, 2 Core(s), and 4 Logical Processor(s), 12 GB RAM hardware configuration.

The curve-fitting model required some value initially for parameters. Initial guesses for parameters are very crucial. We need to know what parameters have known and what we have to get out of it. The proposed model has been used in many parameters. We have computed some parameters and considered some according to the present study and data. We do not need to fit N, just put the population of the place we have to model and similarly no need to calculate Beds_0_, just put the number of ICUs of the place we have to model. We have to give value to some parameters based on real data and analysis, i.e., Social distancing parameter η lies in the range zero and one, where zero indicates everyone is locked down and quarantined while one is for a normal life.

A Chinese study has shown the incubation period to be 5.2 days on average but it varies among different peoples [65]. The Chinese team study found 14 days of medical observation is necessary for those people who are exposed to pathogens.

Median Hospital stay (χ=1/10) and recovery period (γ=1/10)

The JANA study has found that the median hospital stay is 10 days, and all those who discharged alive, and research showed the basic recovery period of the patient is 10 days [66, 67].

The median time to development of pneumonia and other symptoms for hospitalization (Ψ=1/5) and hospital to ICU admission (σ=1/ 7). The median time from symptom onset to the development of pneumonia is approximately 5 days, [68, 69] and the median time from symptom onset to severe hypoxemia and ICU admission is approximately 7–12 days [69-73]. Median intensive care units (ICUs) length of stay (φ =1/8).

The median intensive care units (ICUs) length of stay for COVID-19 patients was approximately 8 to 13 days of respiratory support in a Chinese report[73][74]. Hence the recovery time from critical is the same as ICU’s length of stay that is 8 days (μ=1/8).

The case fatality rate (CFR) represents the proportion of cases who eventually die from the disease The basic formula of case fatality rate is 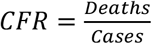 and sometimes this formula is corrected then 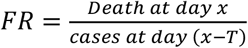, where T = average time from case confirmation to death.

We know that *β*(*t*) can be calculated by basic reproduction number *R_0_*(*t*) and *γ* hence, no need to find any separate parameter for *β*. The beds scaling factor s can be fitted, it not affect much in the result because the amount of people who are treated because of lack of facility is very less as compared to the number of death. The proposed model has three probability estimates *prob*(*I → H), prob*(*H → C*) and *prob*(*C → D*) split up by age groups and weighted by the proportion of per age group. We will try to fit all above probabilities, which extremely close to the prediction obtained of special risk group such as diabetics, high blood pressure, heart disease, etc. now we have to fit all the probabilities *rob*(*I → H*)*, prob*(*H → C*) and *prob*(*C → D*) and 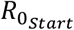, 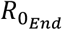 *, x*_0_ and *η* are the parameters of *R*_0_(*t*).

Johns Hopkins University Center for Systems Science and Engineering is the main source of data [60]. We have collected and cleaned data for age groups, probabilities, and ICU beds from UN Data [61].

A total number of Hospital beds and ICUs as per one lack peoples for topmost affected countries. The number of people per age group for topmost affected countries. Probabilities for *prob(I→H), prob(H→C)*, and *prob(C→D)* per age groups but we use fitted probabilities and collect the data of the number of fatalities day wise from 22 January 2020 onwards.

We only use the data of the number of fatalities, not total cases or active cases because the number of reported cases is not much accurate it depends on many factors like the number of test cases, etc, but the number of fatalities reported is more accurate.

We have to collect all the data and parameters first of all that we already know, then define some initial guesses and set the upper and lower bounds for those we don’t know. We can change the upper and lower bound and initial guess according to the situation. We set Levenberg–Marquardt as a fit model after defining all parameters. Levenberg–Marquardt algorithm is the most important algorithm in data fitting [62, 63]. Lmfit library provides a high-level interface to non-linear optimization and curve-fitting problems for Python [64]. R-squared value is a measure of how close the data are to the fitted line. R-squared value is always between zero and one. In general, the higher the R-squared value, the better the model fits your data. Curve fitting of death cases of India and Brazil is shown in Fig. 6.

**Table 1:**
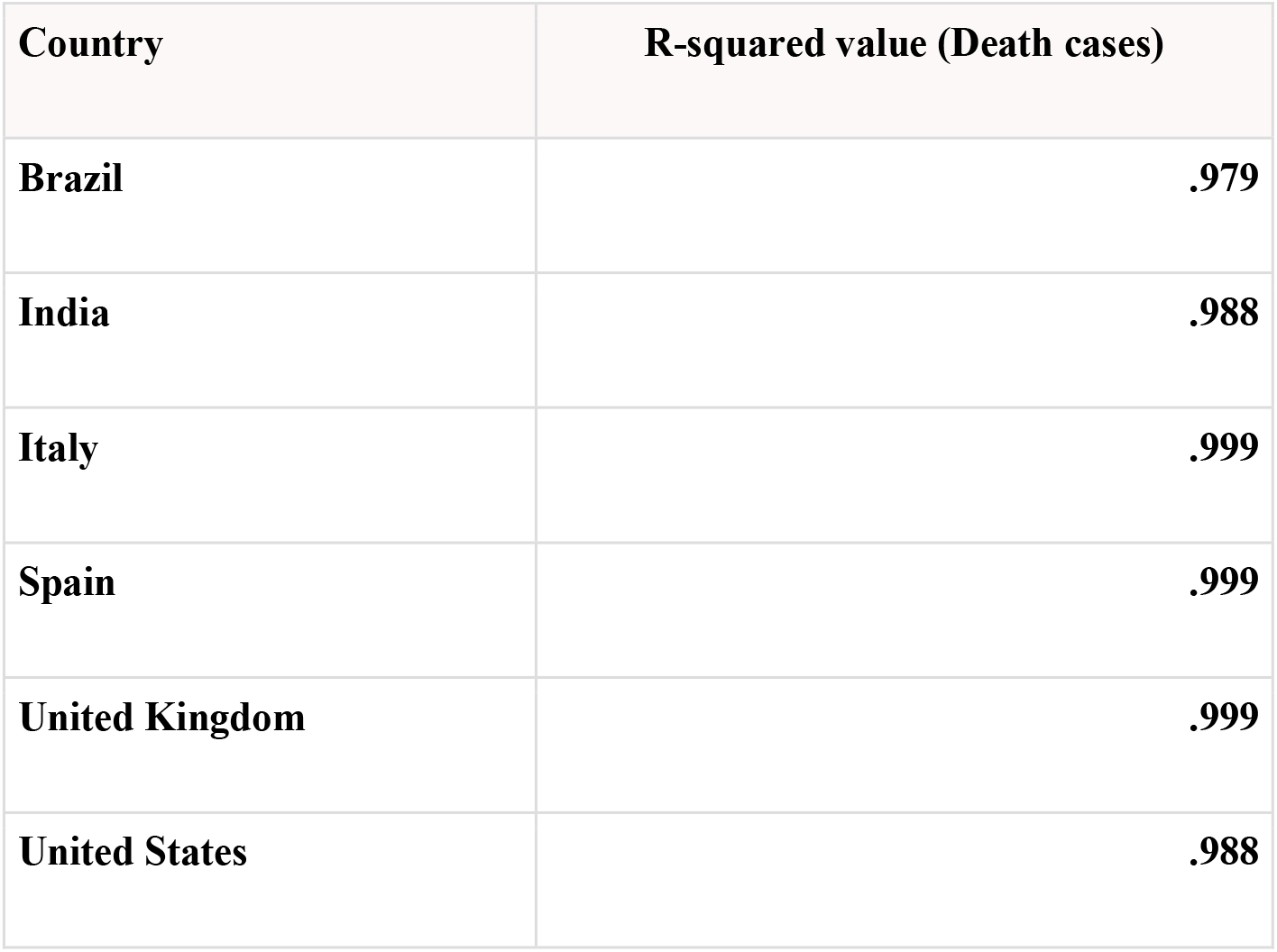
R-squared value of curve fitting for Brazil, India, Italy, Spain, United Kingdom, and the United States

**Figure 6:**
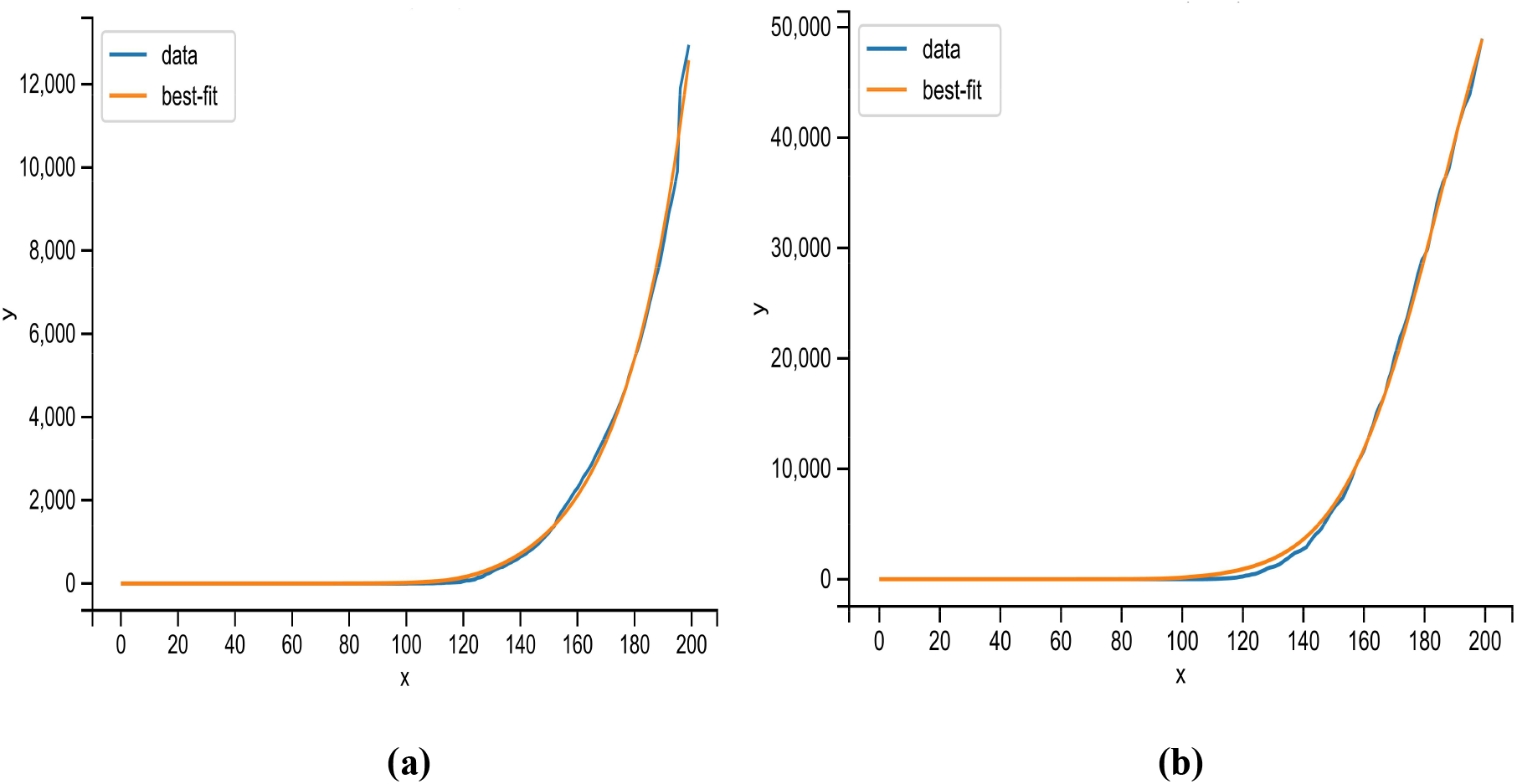
Curve fitting of death cases of India and Brazil

A Chinese study has found, 80% of cases are mild. Based on all 72,314 cases of COVID-19 confirmed, suspected, and asymptomatic cases in China as of February 11, a paper by the Chinese CCDC released on February 17 and published in the Chinese Journal of Epidemiology[76-80] has found that:

1. 80.9% of infections are mild and can recover at home.
2. 13.8% are severe, developing severe diseases including pneumonia and shortness of breath.
3. 4.7% as critical and can include respiratory failure, septic shock, and multi-organ failure.
4. in about 2% of reported cases, the virus is fatal.
5. The risk of death increases the older you are.
6. Relatively few cases are seen among children.

Pre-existing illnesses that put patients at higher risk:

1. cardiovascular disease
2. diabetes
3. chronic respiratory disease
4. hypertension

We have collected and analyzed data from all US States. In the meantime, below we have shown the data provided by New York City Health as of May 13, 2020 [81, 64]:

**Table 2:** Age-wise rate of hospitalization from infected cases, critical cases from infected cases, and deceased cases from ICUs

| AGE | Rate of hospitalization from Infected cases | Rate of critical cases from Infected cases | Rate of deceased cases from ICUs |
| --- | --- | --- | --- |
| 80+ years old | 18.4% | 0.16% | 0.98% |
| 70-79 years old | 16.6% | 0.10% | 0.86% |
| 60-69 years old | 11.8% | 0.07% | 0.57% |
| 50-59 years old | 8.2% | 0.05% | 0.26% |
| 40-49 years old | 4.3% | 0.03% | 0.10% |
| 30-39 years old | 3.4% | 0.025% | 0.06% |
| 20-29 years old | 1.0% | 0.009% | 0.05% |
| 10-19 years old | 0.1% | 0.003% | 0% |
| 0-9 years old | 0.01% | 0.001% | 0% |

### COVID-19 Fatality Rate by AGE

We know Death Rat**e** = (number of deaths/number of cases), can be written as the probability of dying if infected by the virus (in percentage) and it depends on the different age groups. In Tab. 3 percentages represent, for a person in a given age group, the risk of dying if infected with COVID-19 [82].

**Table 3:** Age-wise death rate of all cases and comorbidity cases

| Age | Death rate of all cases | The death rate of comorbidity cases |
| --- | --- | --- |
| 80+ years old | 14.8% | 0.98% |
| 70-79 years old | 8.0% | 0.86% |
| 60-69 years old | 3.6% | 0.57% |
| 50-59 years old | 1.3% | 0.26% |
| 40-49 years old | 0.4% | 0.10% |
| 30-39 years old | 0.2% | 0.06% |
| 20-29 years old | 0.2% | 0.05% |
| 10-19 years old | 0.2% | 0% |
| 0-9 years old | ( no fatalities) 0% | 0% |

### COVID-19 Fatality Rate by COMORBIDITY

The comorbidity fatality rate for all the cases has shown in Tab. 4 and the percentage showed that for a patient with a given pre-existing condition, the risk of dying if infected by COVID-19 [83].

**Table 4:** For all top comorbidity cases death rate

| PRE-EXISTING CONDITION | DEATH RATE all cases |
| --- | --- |
| Cardiovascular disease | 10.5% |
| Diabetes | 7.3% |
| Chronic respiratory disease | 6.3% |
| Hypertension | 6.0% |
| Cancer | 5.6% |
| No pre-existing conditions | 0.9% |

We have used the publicly available dataset of COVID-19 provided by the Johns Hopkins University [84]. This dataset includes many countries’ daily count of confirmed cases, recovered cases, and deaths. This time series dataset is available from 22 January 2020. We also gathered and crosschecked data in Worldometer, Coronavirus cases [81], a website providing real-time data of COVID-19. These data are collected through public health authorities’ announcements and are directly reported public and unidentified patient data, hence ethical approval is not required.

### Distribution of Total cases worldwide

WHO has reported that 216 countries are affected by COVID-19. Fig. 7 shows that 35% of cases are from six countries only. The United States of America has the highest number of cases, which alone is 19% of the world.

**Figure 7:**
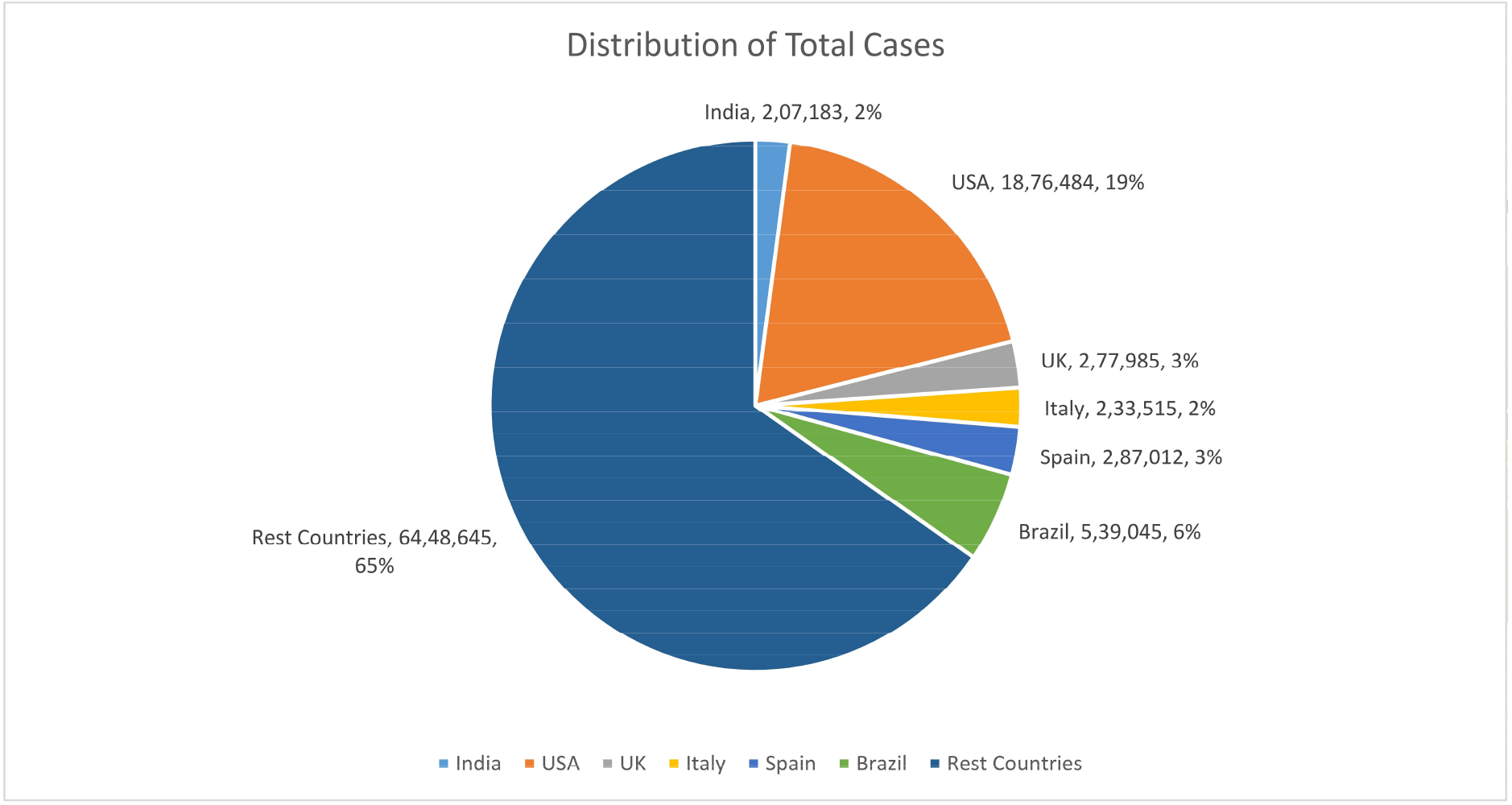
Distribution of total cases worldwide

### Distribution of Death cases worldwide

Death cases are more accurate because rarely any case from COVID-19, which has not been registered. Fig. 8 shows that 39% of cases are from six countries only. The United States of America has the highest number of cases, which alone is 17% of the world.

**Figure 8:**
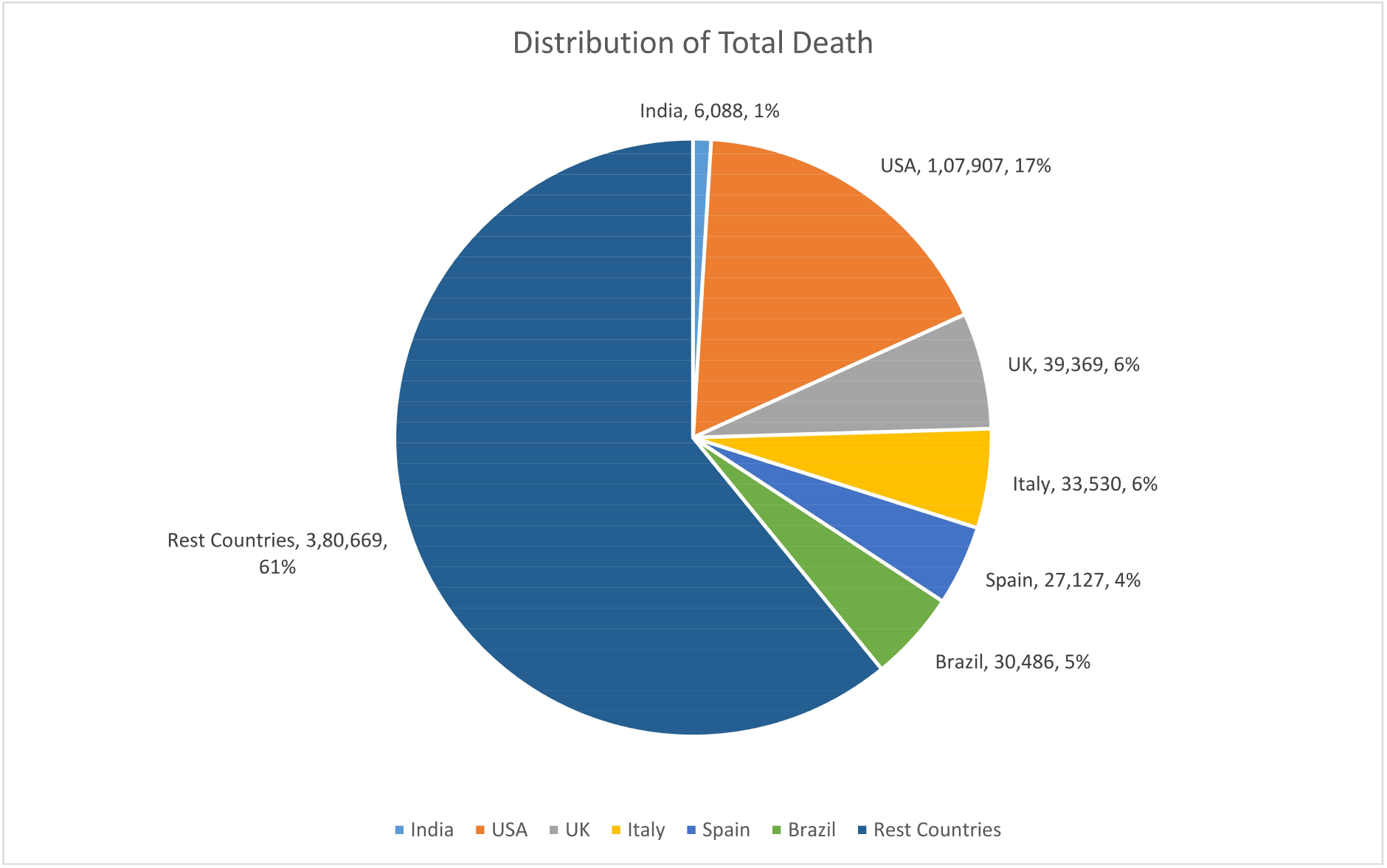
Distribution of Death cases worldwide **Total cases:** The fig. 9 shows the Country wise total cases from 22 January 2020 to 20 June 2020 of COVID-19.

**Figure 9:**
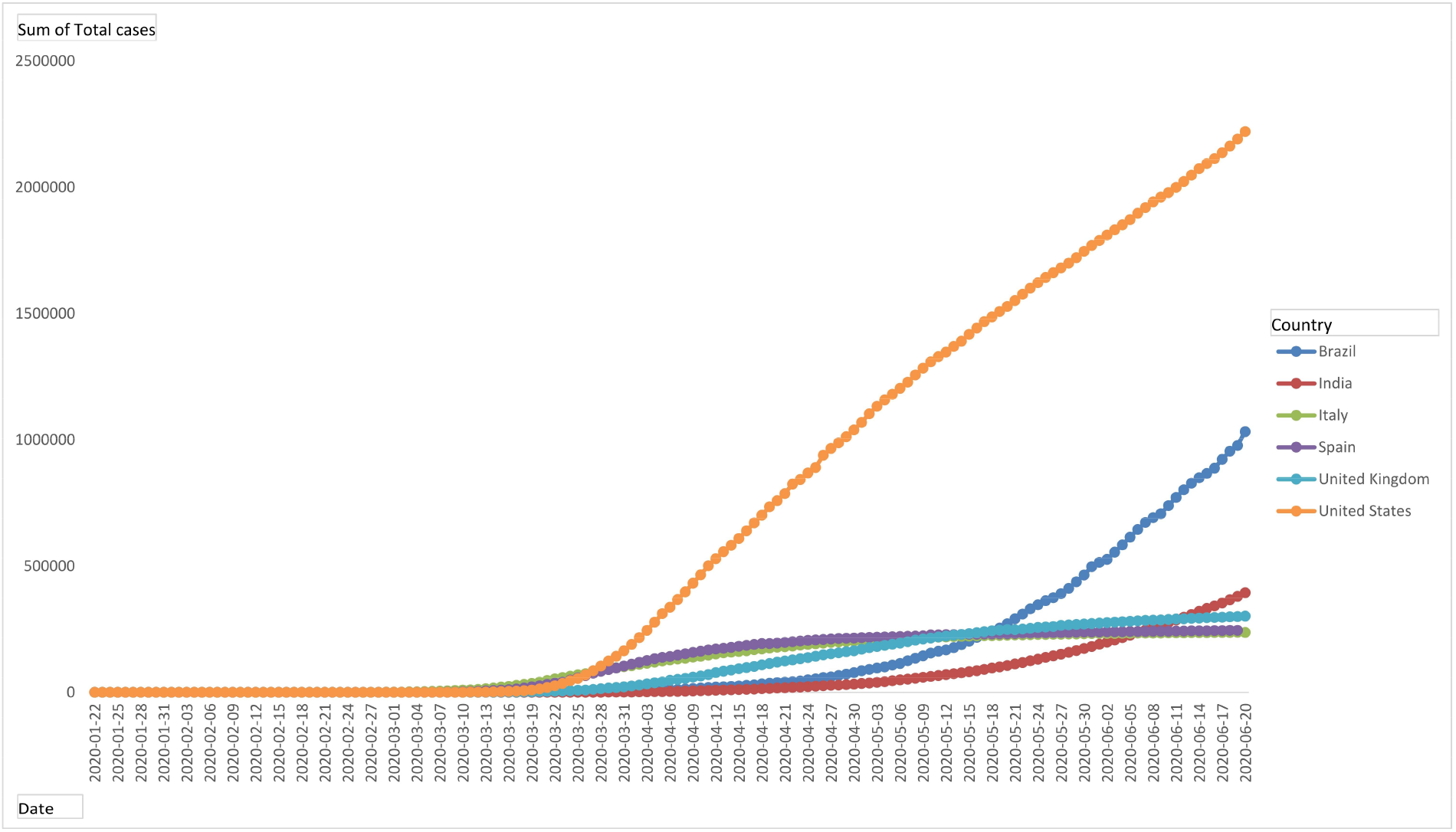
Day-wise total cases of Brazil, India, Italy, Spain, United Kingdom, and the United States **Total death:** The fig. 10 shows the Country wise total death from 22 January 2020 to 20 June 2020 of COVID-19.

**Figure 10:**
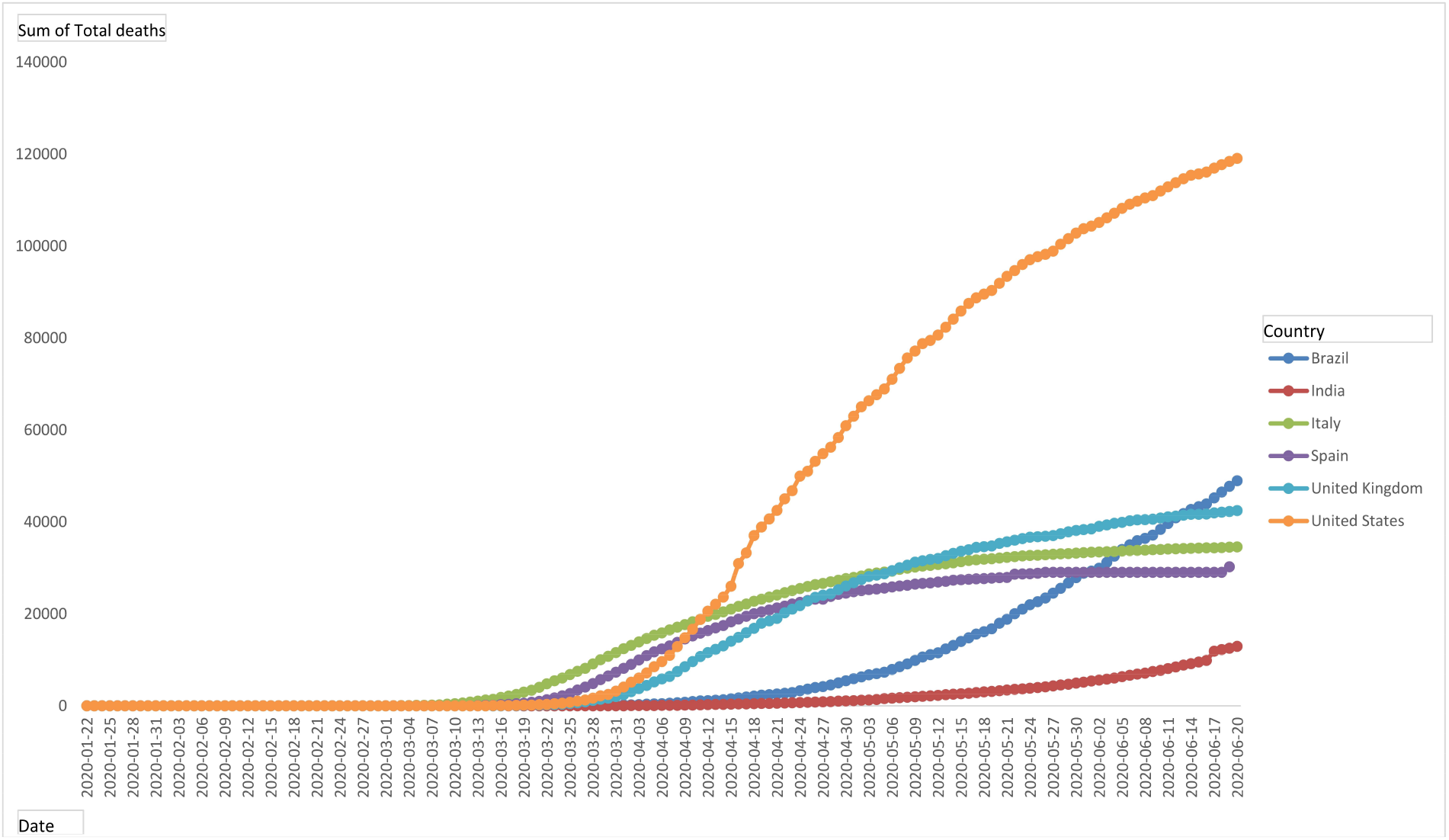
Day-wise total death of Brazil, India, Italy, Spain, United Kingdom, and the United States **Daily new cases:** The fig. 11 shows the Country wise daily new cases from 22 January 2020 to 20 June 2020 of COVID-19.

**Figure 11:**
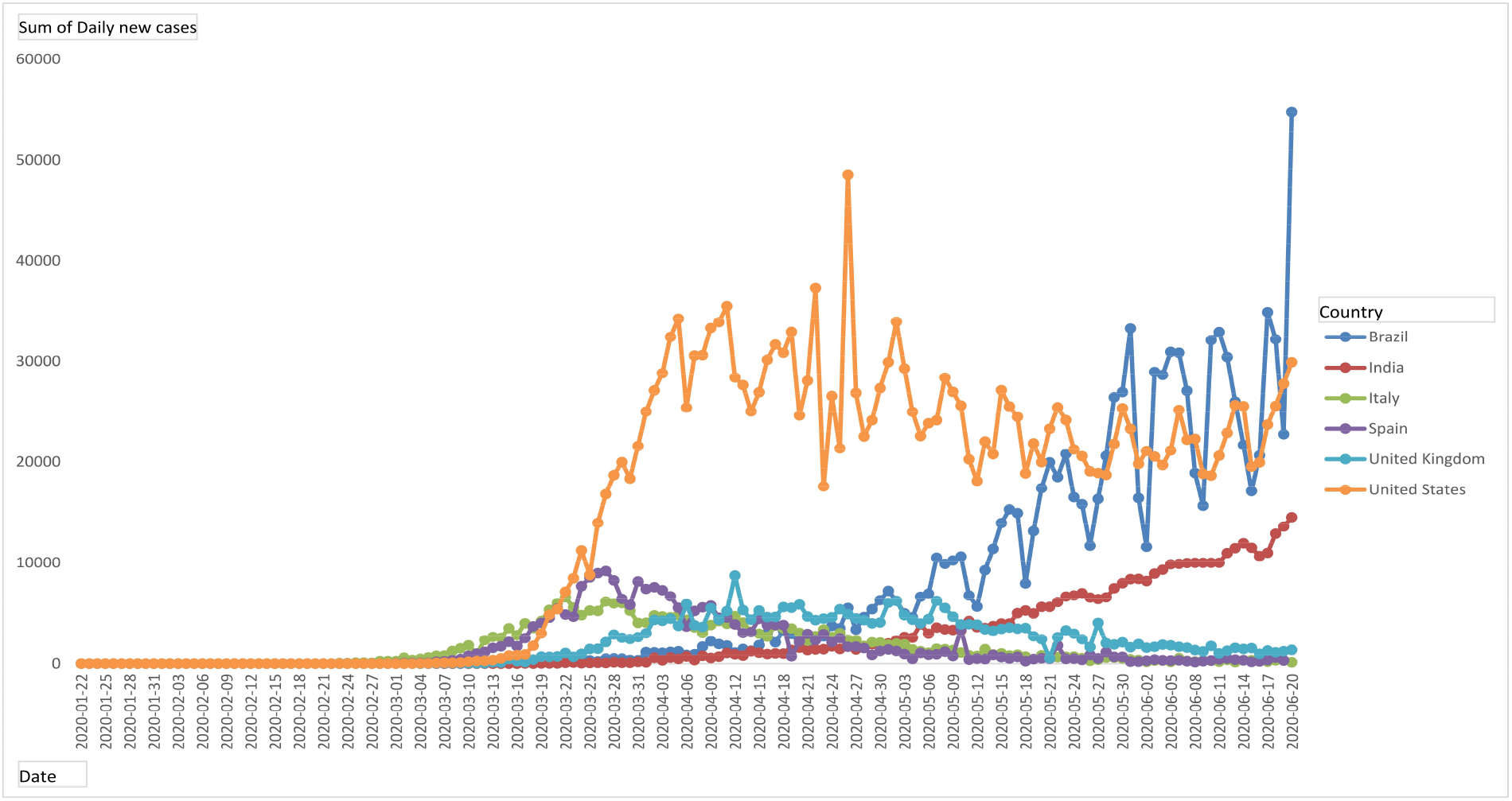
Daily new cases of Brazil, India, Italy, Spain, United Kingdom, and the United States **Daily new death:** The fig. 12 shows the Country wise daily new death cases from 22 January 2020 to 20 June 2020 of COVID-19.

**Figure 12:**
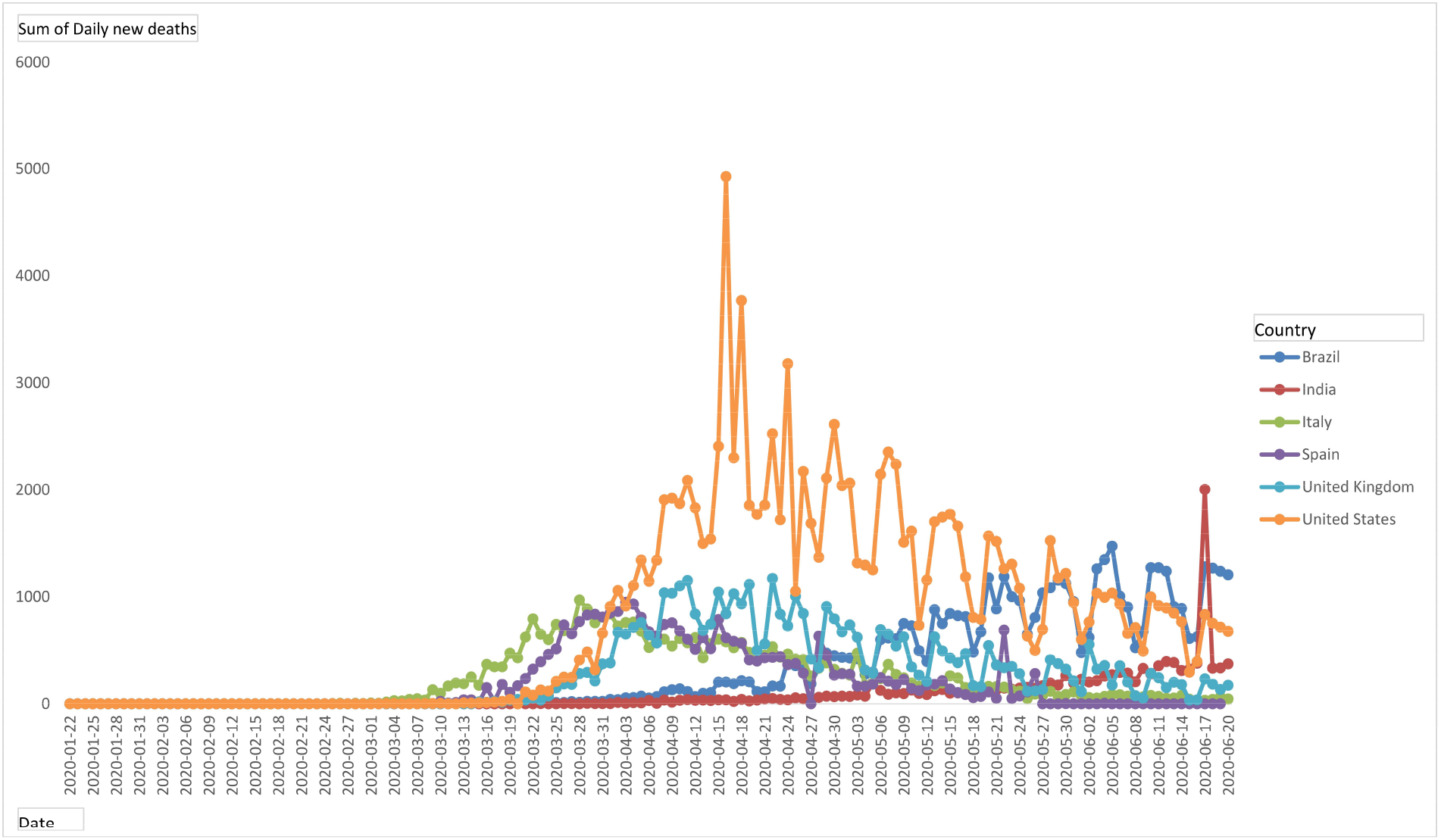
Daily new death cases of Brazil, India, Italy, Spain, United Kingdom, and the United States **Case fatality rate:** It is difficult to estimate the overall case fatality rate of an ongoing pandemic but we can calculate the daily fatality rate. Fig. 13 shows the Country wise daily case fatality rate from 01 March to 03 June 2020 of COVID-19.

**Figure 13:**
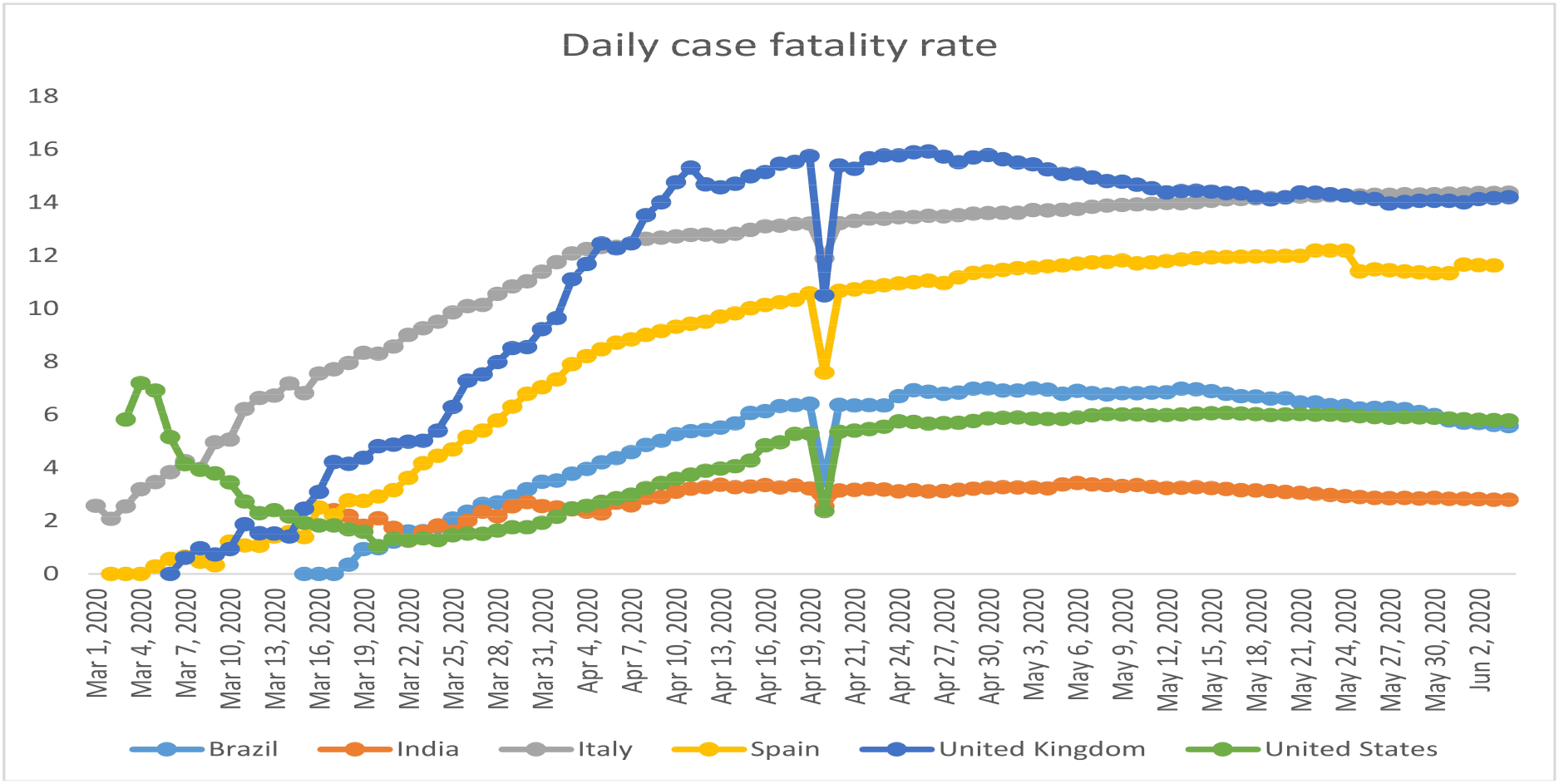
Daily case fatality rate of Brazil, India, Italy, Spain, United Kingdom, and the United States

### Age-wise population

Italy and Spain in the 70 plus year-wise age category population is more percentage-wise, according to World Bank data. India and Brazil’s population is higher in the age category of o to 20 years. Brazil, India, Italy, Spain, the United Kingdom, and the United States, the distribution of the population are almost equal between 20 and 70 age groups in all these countries. The population has distributed according to age category is shown in fig. 14.

**Figure 14:**
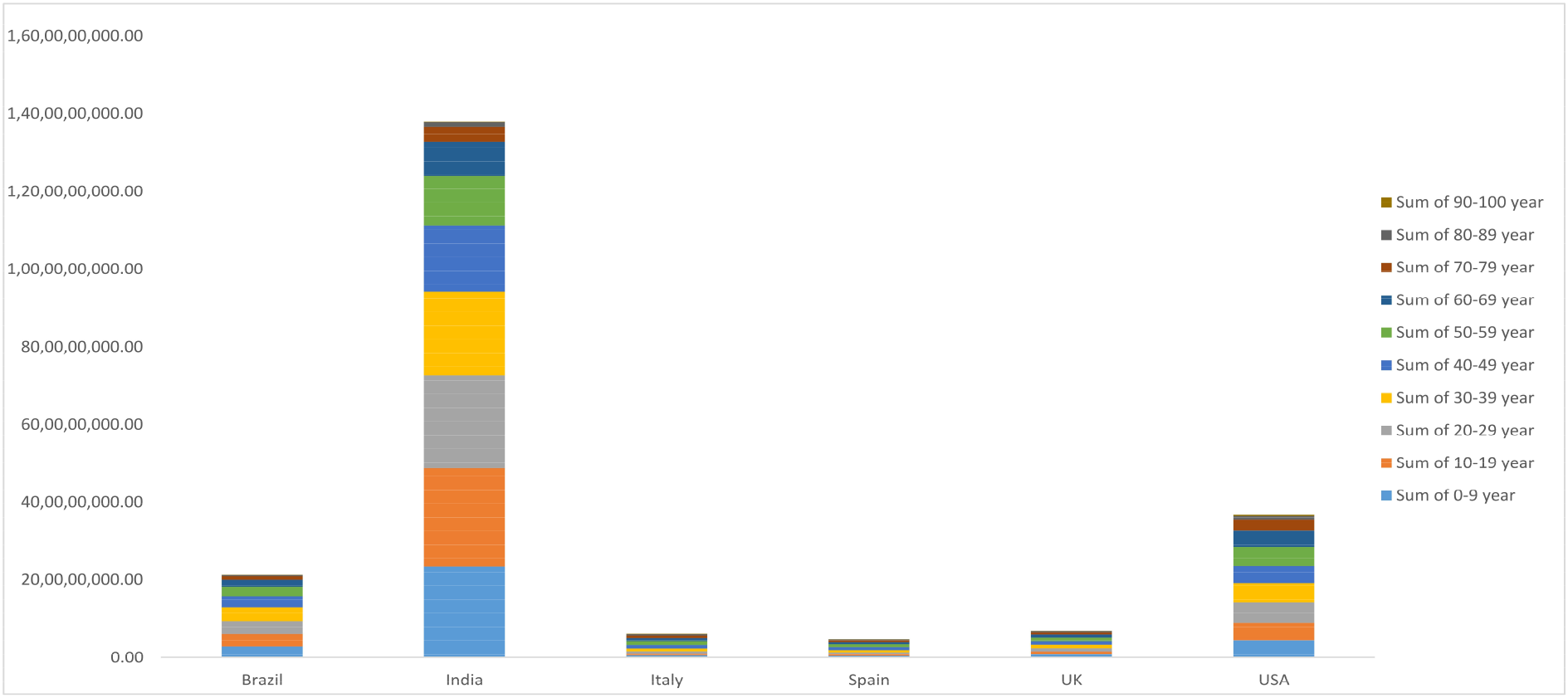
The distribution of the population according to the age category of Brazil, India, Italy, Spain, United Kingdom, and the United States

### Number of hospital beds per 100k

Hospital beds or curative beds counting unit beds per 100,000 population or 100k population according to WHO and OECD. The basic measures focus on all the hospital beds, which are occupied or empty. The Country-wise number of hospital beds has shown in Fig. 15.

**Figure 15:**
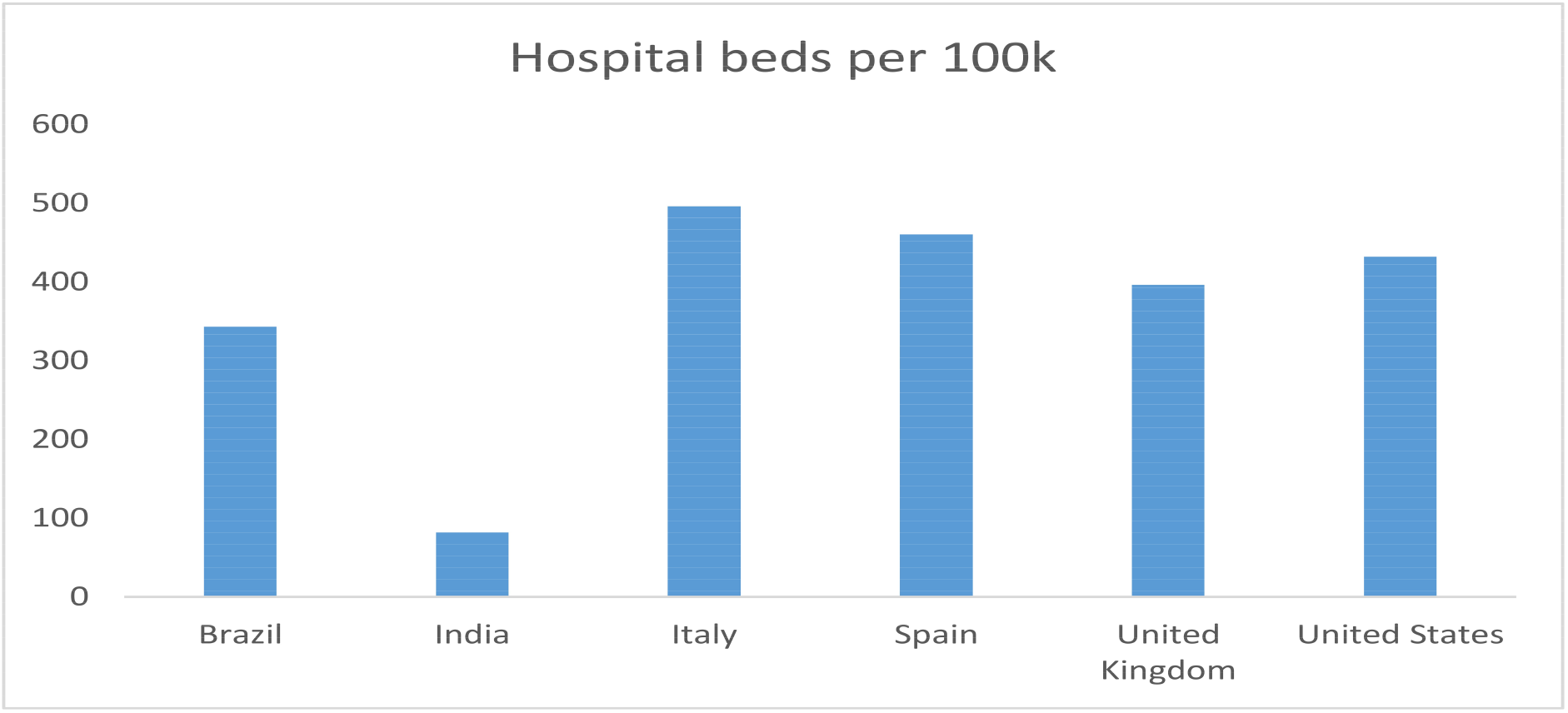
The Country-wise number of hospital beds of Brazil, India, Italy, Spain, United Kingdom, and the United States

### Number of ICU beds per 100k

ICU beds counting unit is beds per 100,000 population or 100k population according to WHO and OECD. ICU beds have provided to all the cases with severe conditions. The Country-wise number of ICU beds is shown in Fig. 16.

**Figure 16:**
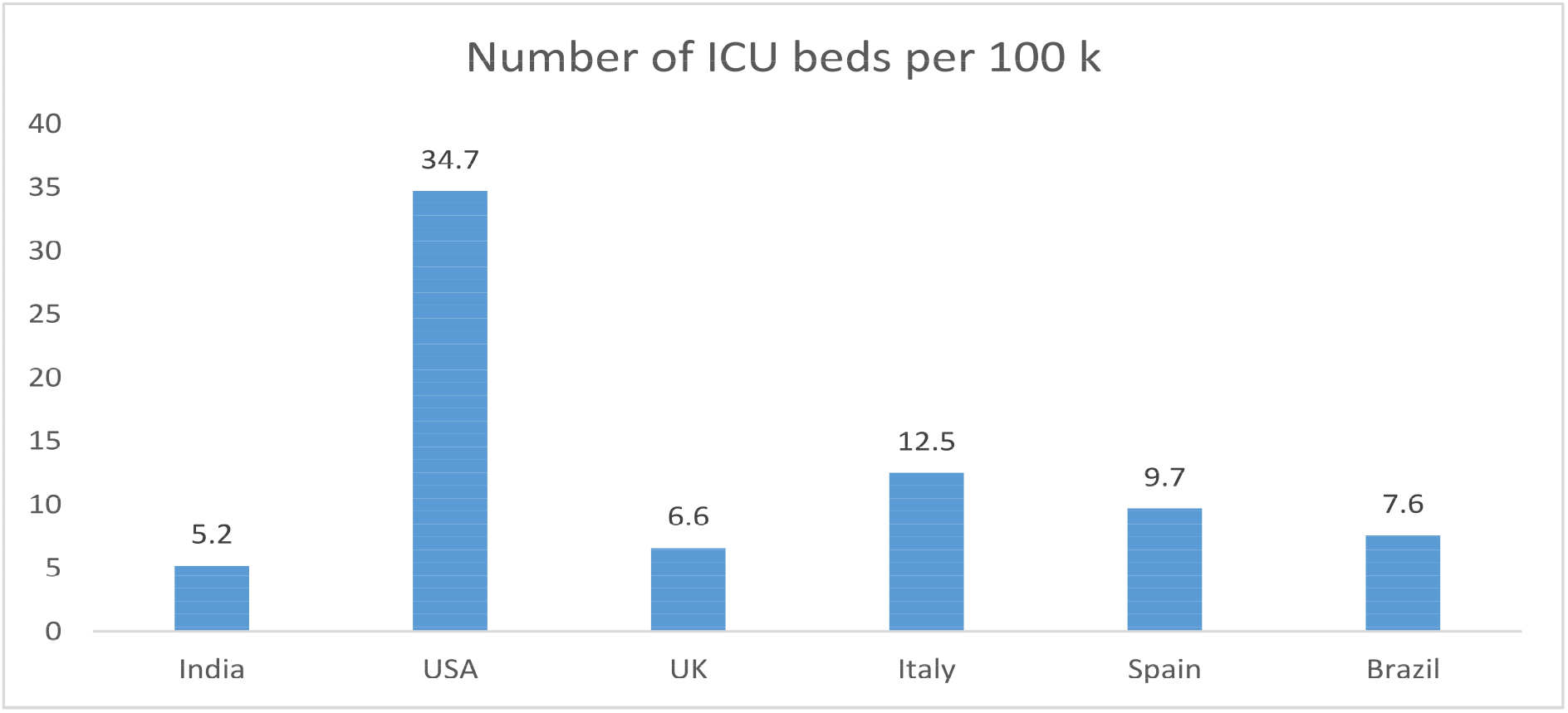
The Country-wise number of ICU beds of Brazil, India, Italy, Spain, United Kingdom, and the United States

## 4 Results

We have analyzed and predict the COVID-19 trend among the top affected countries of the world using our proposed SEIHCRD model in this section. Our proposed methods outcome shows Italy, Spain, the United Kingdom, the United stated of America have seen their worst time, now their conditions are improving. The peak of Brazil and India is yet to come in June and July month. Brazil will need hospital beds and ICU beds when the number of cases will become very high in the middle and last of June. Disease distribution in India is very random, e.g. Number of cases in Maharashtra and Delhi are very high in comparison to other states, some beds needed in June especially in Delhi state. Our method shows that Italy already suffers its peak now they shuttle down. The proposed model has shown that endpoint of USA cases in September, and UK cases will shuttle down in July. We have compared our proposed method results with real-world data, then it is largely the same.

### 4.1 SEIHCRD Model for Brazil

Brazil has the second-highest in COVID-19 cases just behind the USA as of the starting of June 2020. COVID-19 cases will be at a peak in Brazil according to our proposed method in June and July. Basic reproduction number in nearly 4.5 in initial days, after some days it decreases, and after 220 days, it becomes one or even less. The case fatality rate has ups and down daily but it goes higher and higher up to 7.5 in our model and then slow down and total CFR is nearly 3.0 overall. The result of our proposed method shows that in Brazil, the number of cases will increase hence Brazil will need some Hospital beds and ICU beds in June, July, and August. A daily number of death cases on top of nearly 190^th^ day and goes up to 1800 people die per day, then slow down and shuttle down in October. The model has shown nearly 15 to 20 days, 700 hospital beds and 400 ICU beds required on-peak time. SEIHCRD Model results for Brazil has shown in Fig. 17.

**Figure 17:**
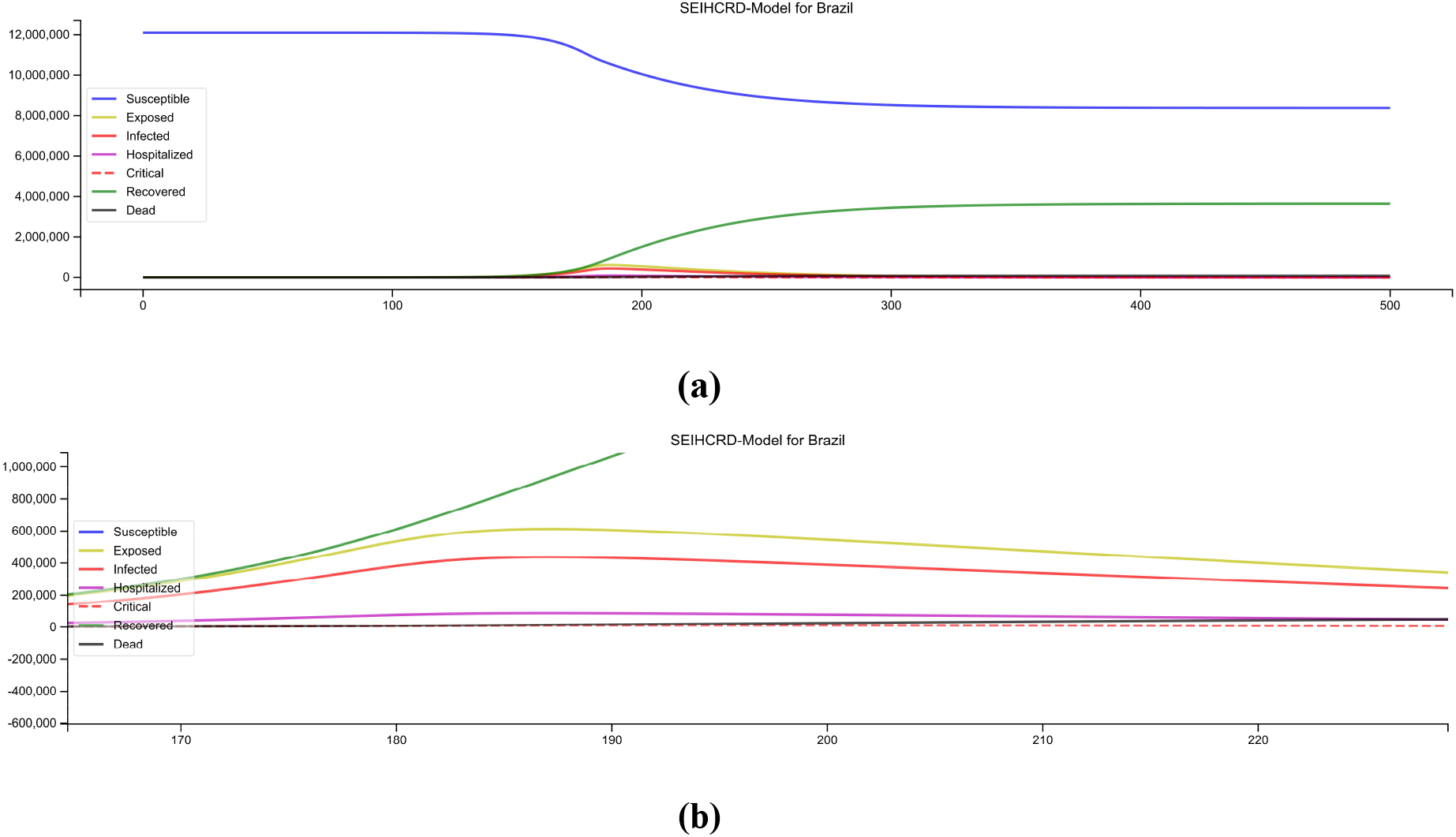
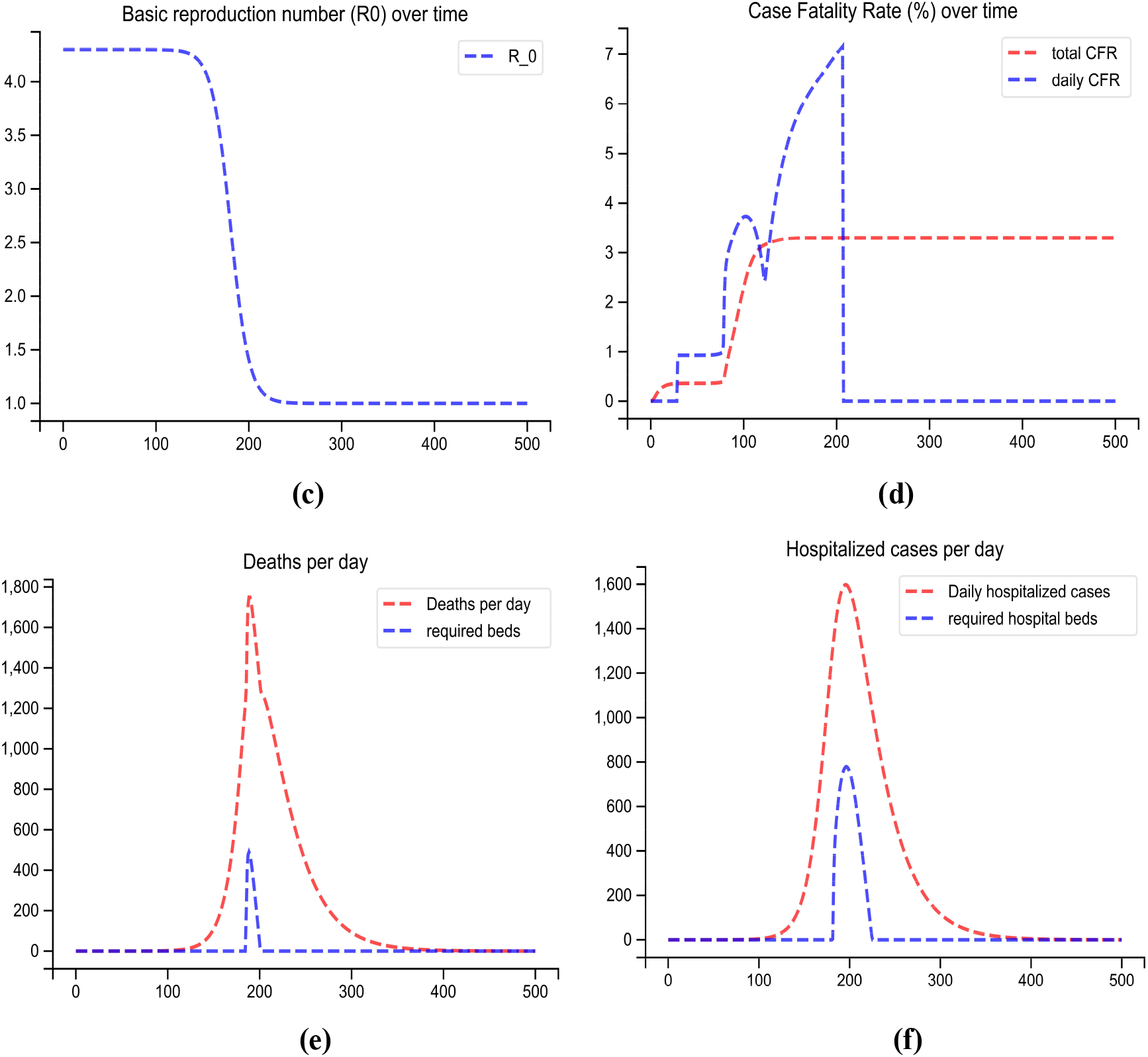
SEIHCRD Model for Brazil (a) Spread scenario of SEIHCRD Model for Brazil. A 500-day analysis has been done by the proposed model, which starts from 22 January. (b) In this, the peak point of the model has shown by zooming for cases. (c) The basic reproduction number of Brazil over time (d) The case fatality rate of Brazil over time. (e) Redline shows the number of deaths per day and the blue line shows how much ICU beds are required in peak days. (f) Redline shows the number of cases hospitalized per day and the blue line shows how much hospital beds are required in peak days.

### 4.2 SEIHCRD Model for India

The SEIHCRD model results show COVID-19 cases are on the peak in India around 200^th^ day (June last or July month) and there will no cases come at the end of September and October month. SEIHCRD-Model for India shows that the case fatality rate of COVID-19 in India is very low. Basic reproduction number R_0_ is nearly 2.0 in starting, it decreases after some time and falls slowly and after 170 days it is 1.3 that is nearly equal to the real scenario, and it goes down than one or even lower in the next 50 to 60 days. The daily Case fatality rate varies between 1.5, 3.0, and the overall case-fatality rate very steady after 200 days of nearly 2.0. India’s geographical condition is completely different from the rest of the countries. The spread of COVID-19 disease is random in India. Maharashtra, Delhi Tamil Nadu, and Gujrat are most disease-affected states. According to the proposed model, June last and July starting the number of patients admitted to the hospital on the peak. Hospital beds and ICU beds have needed in June in India especially in Delhi states. Around the 200^th^ day when the cases are at peak then the number of death cases reaches 550 and the need of nearly 250 Hospital beds and ICU beds. SEIHCRD Model results for India are shown in Fig. 18.

**Figure 18:**
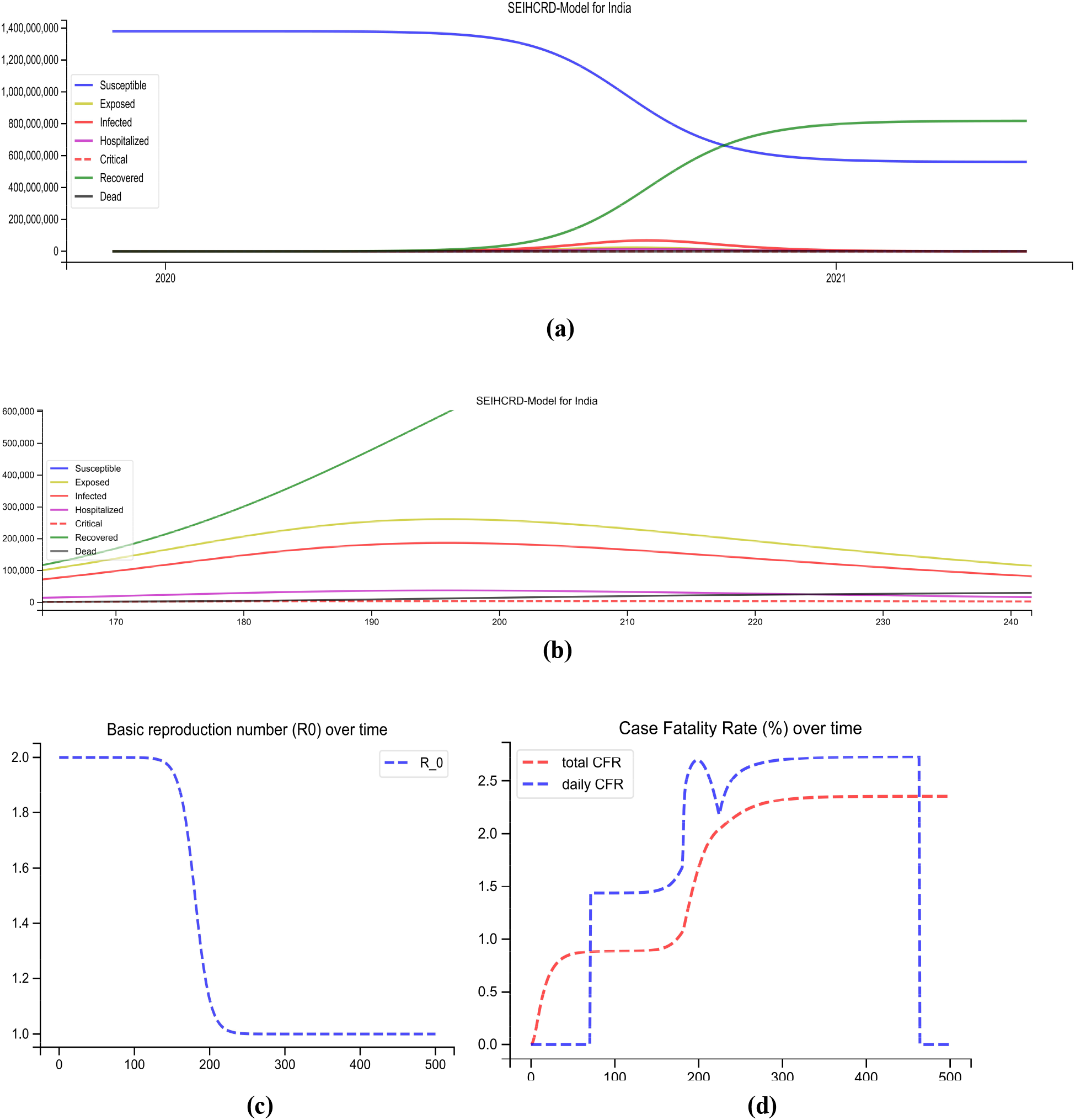
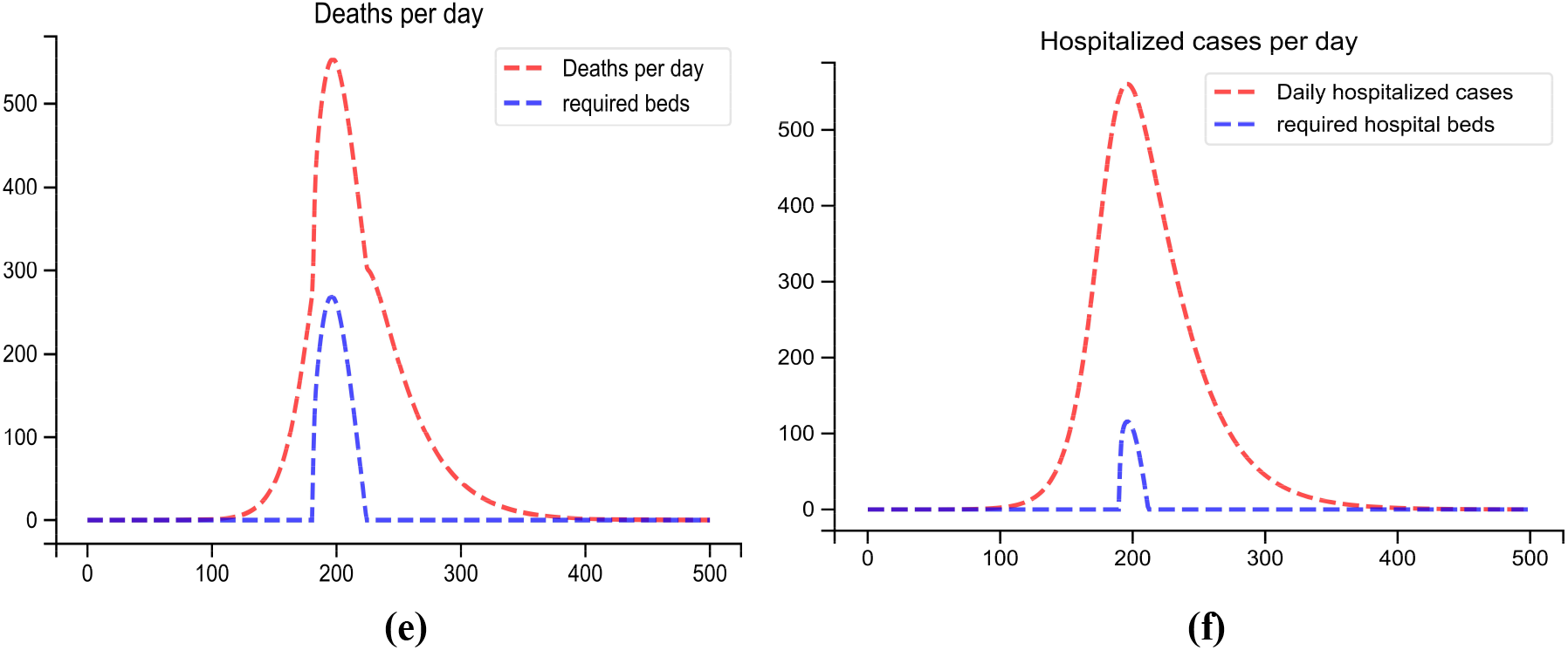
SEIHCRD Model for India (a) Spread scenario of SEIHCRD Model for India. A 500-day analysis has been done by the proposed model, which starts from 22 January. (b) In this, the peak point of the model is shown by zooming for cases. (c) The basic reproduction number of India over time (d) The case fatality rate of India over time. (e) Redline shows the number of deaths per day and the blue line shows how much ICU beds are required in peak days. (f) Redline shows the number of cases hospitalized per day and the blue line shows how much hospital beds are required in peak days.

### 4.3 SEIHCRD Model for Italy

SEIHCRD model result shows, Italy has seen its bad phase and now in a state of improvement and is trying to recover from that disastrous pandemic. Italy’s healthcare system is the second-best in the world according to WHO. COVID-19 cases on a peak between 70 and 90^th^ days in Italy, and after 220 days its cases are very rare, and its cases almost stop coming in August. Basic reproduction number R_0_ starts from 5.0 and after about 100 days, the value of R_0_ starts decreasing visually. The number of death decrease drastically in the next few days after peak days. The proposed model for Italy showing that around 100^th^ day, the number of hospital beds and the number of ICU beds are required. That time the number of death cases is 800 per day and daily-hospitalized cases are nearly 2500. The Case fatality rate value reached 7.0, after that its value is reduced drastically in Italy. SEIHCRD Model results for Italy has shown in Fig. 19.

**Figure 19:**
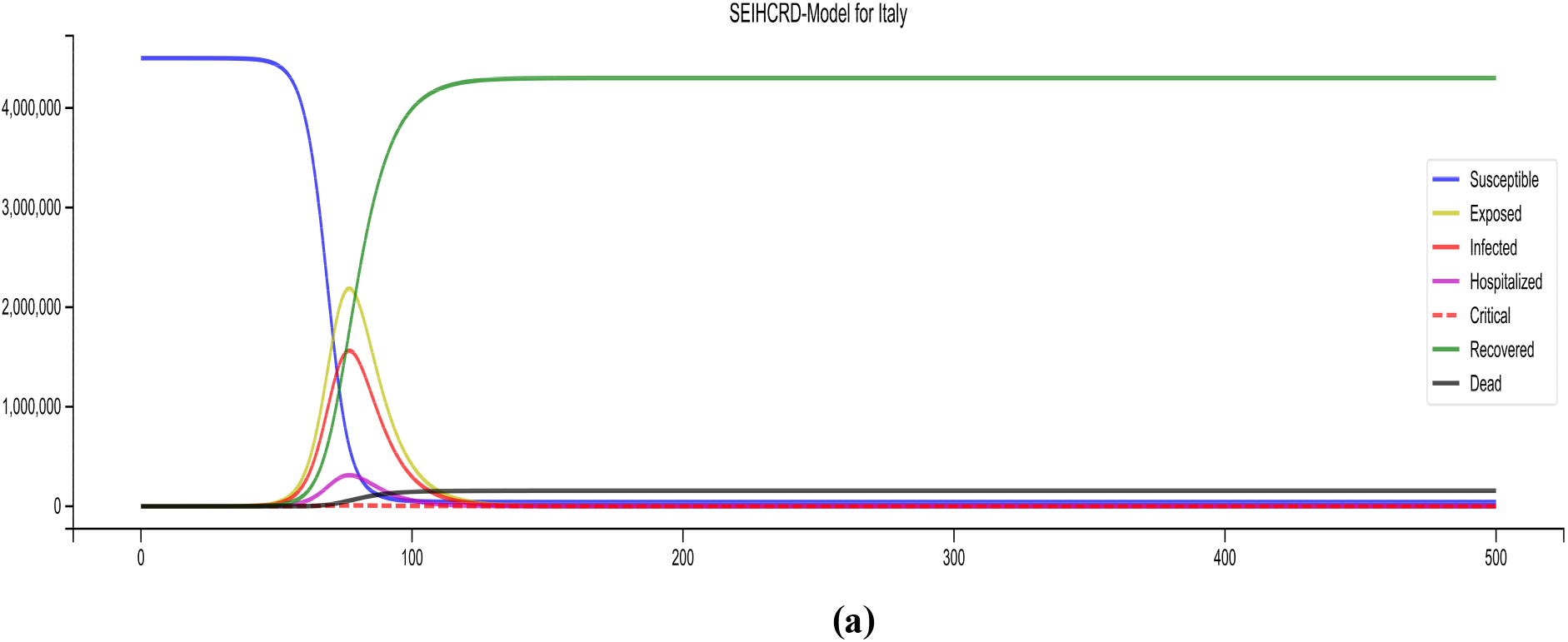
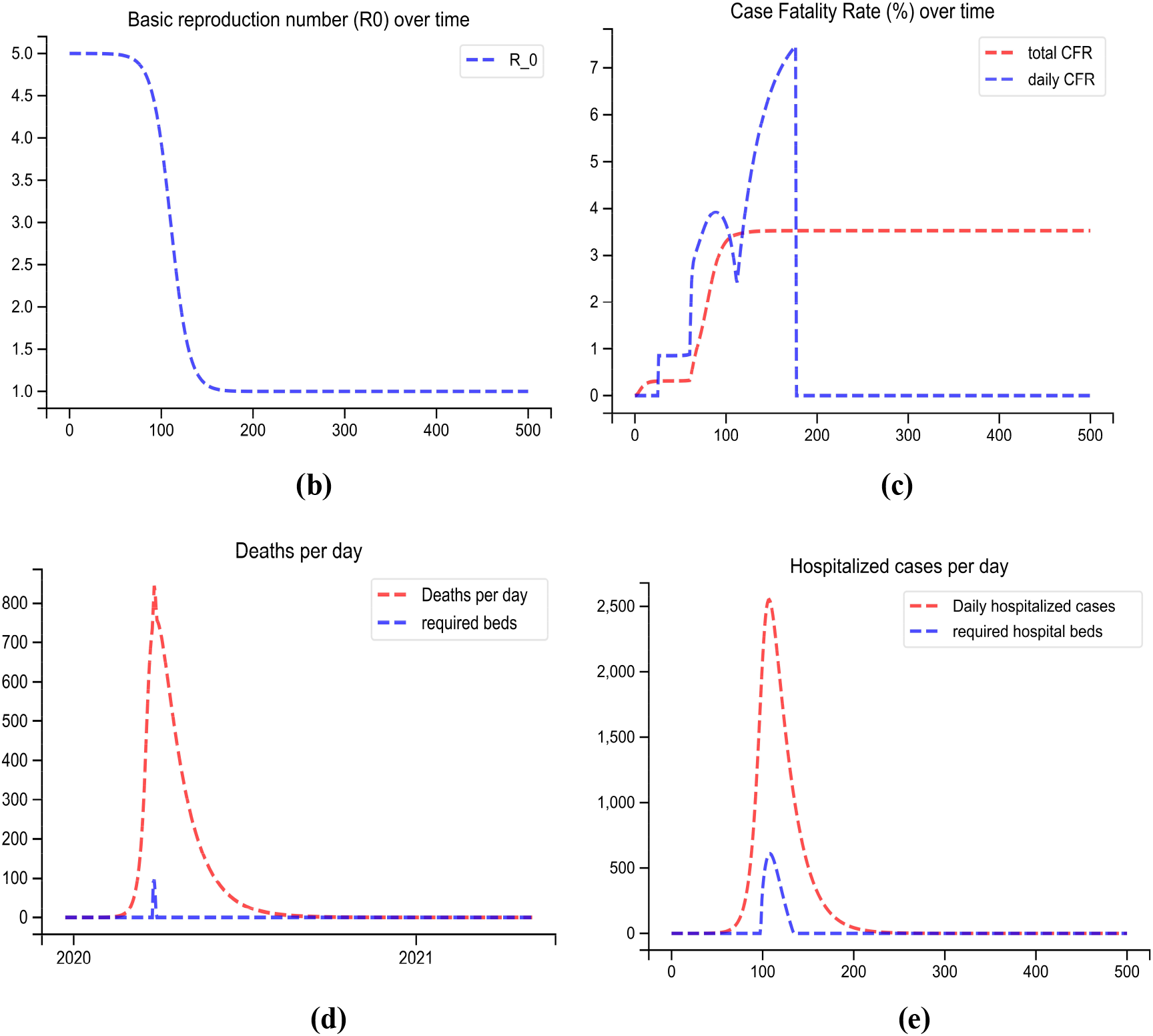
SEIHCRD Model for Italy (a) Spread scenario of SEIHCRD Model for Italy. A 500-day analysis has been done by the proposed model, which starts from 22 January (b) The basic reproduction number of Italy over time (c) The case fatality rate of Italy over time. (d) Redline shows the number of deaths per day and the blue line shows how much ICU beds are required in peak days. (e) Redline shows the number of cases hospitalized per day and the blue line shows how much hospital beds are required in peak days.

### 4.4 SEIHCRD Model for Spain

Spain has also gone through its worst phase. COVID-19 cases are on the peak in Spain around the 110^th^ day according to the SEIHCRD model, and after that, Spain recovered by this disaster rapidly in the next few days. The result shows that COVID-19 cases will not come in June last or July. The basic reproduction number is 7.0, after some time it reduces, and around 170^th^ day it goes to one and below. The Case fatality rate goes up to 7.0, after that its value reduced drastically and it gets one and below around 170^th^ day. In the proposed model for Spain showing that between 110 and 130^th^ days some hospital beds and ICU beds are required. At that, time the number of deaths is nearly 700 per day and daily-hospitalized cases are nearly 1250. SEIHCRD Model results for Spain has shown in Fig. 20.

**Figure 20:**
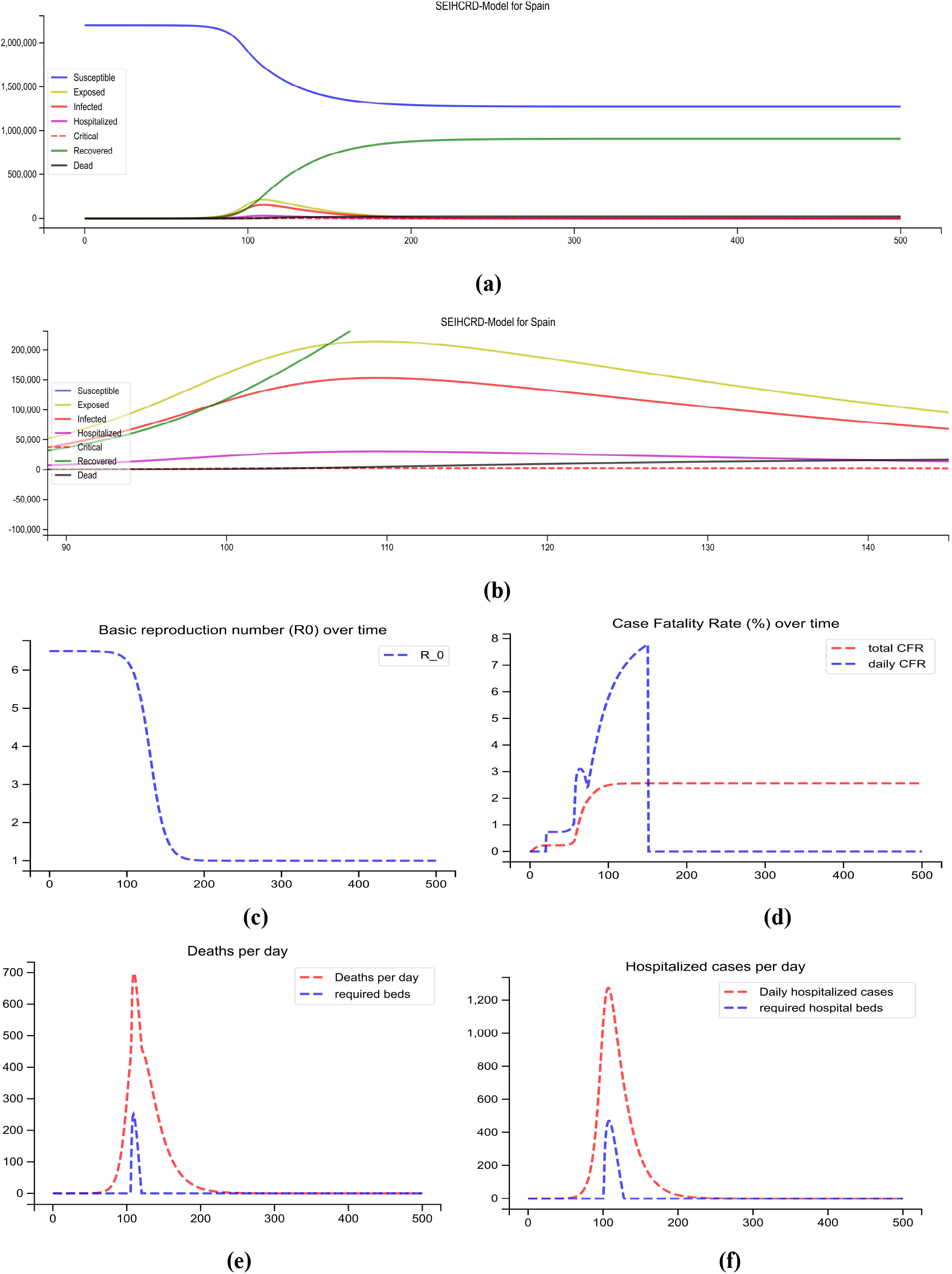
SEIHCRD Model for Spain (a) Spread scenario of SEIHCRD Model for Spain. A 500-day analysis has been done by the proposed model, which starts from 22 January. (b) In this, the peak point of the model is shown by zooming for cases. (c) The basic reproduction number of Spain over time (d) The case fatality rate of Spain over time. (e) Redline shows the number of deaths per day and the blue line shows how much ICU beds are required in peak days. (f) Redline shows the number of cases hospitalized per day and the blue line shows how much hospital beds are required in peak days.

### 4.5 SEIHCRD Model for the United Kingdom

SEIHCRD-Model for the United Kingdom shows that COVID-19 cases on peak around 140^th^ day and after that the United Kingdom has recovered by this disaster very fast in the next few days. After 240 days, its cases are very rare, and its cases almost stop in August. Basic reproduction number R_0_ starts from 4.5 and after about 120^th^ day, the value of R_0_ starts decreasing visually. There are many ups and downs in the daily case fatality rate but it goes higher and higher up to 7.5 in our model and then slows down and total CFR is nearly 3.0 overall. SEIHCRD model has shown for the United Kingdom that some hospital beds and ICU beds have been required between the 100^th^ and 120^th^ days. That time the number of death is 1100 per day and daily-hospitalized cases are nearly 800. The population of old age people is more in the United Kingdom; it has been shown in the previous section. SEIHCRD Model results for the United Kingdom has shown in Fig. 22.

**Figure 22:**
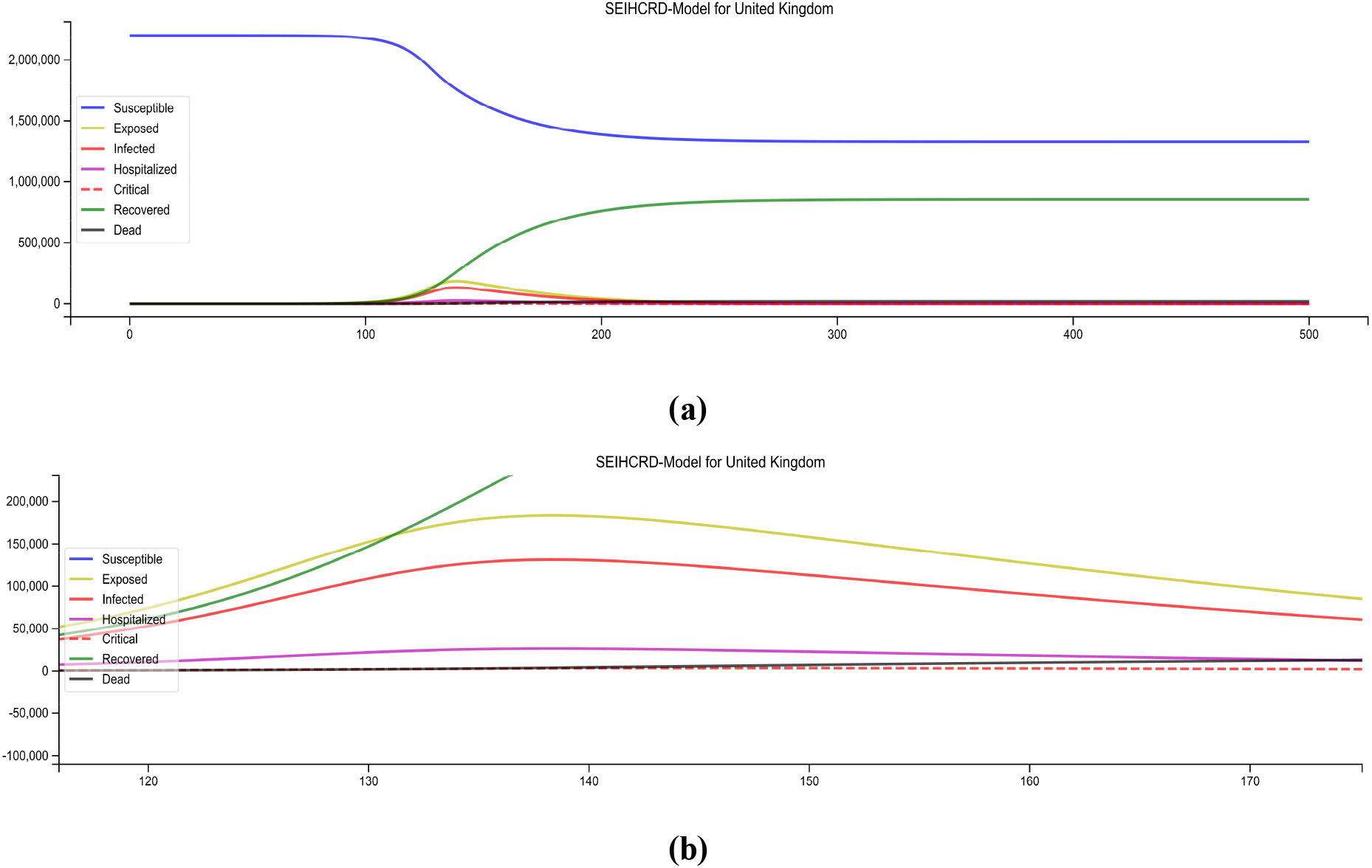
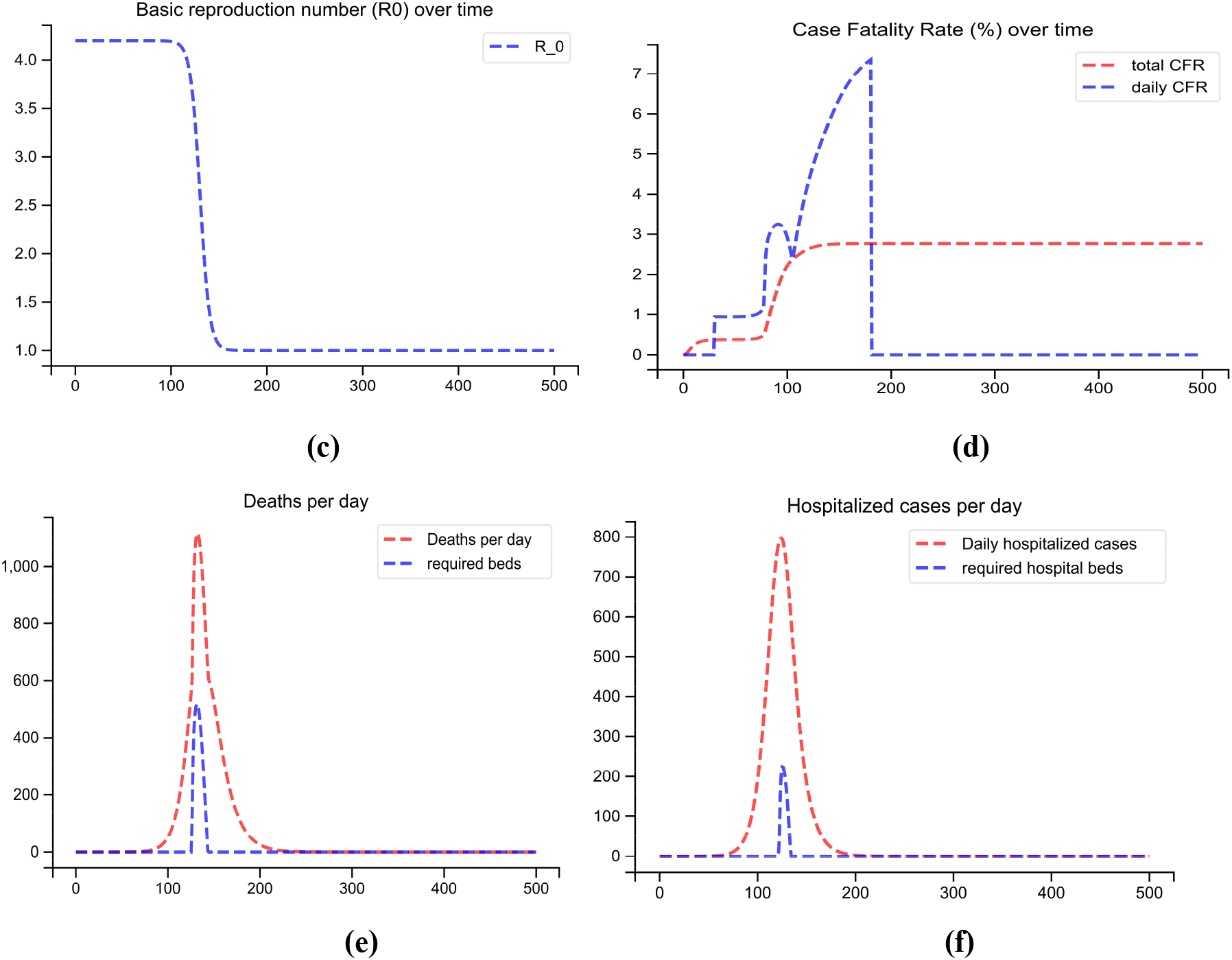
SEIHCRD Model for the United Kingdom (a) Spread scenario of SEIHCRD Model for the United Kingdom. A 500-day analysis has been done by the proposed model, which starts from 22 January. (b) In this, the peak point of the model is shown by zooming for cases. (c) The basic reproduction number of the United Kingdom over time (d) The case fatality rate of the United Kingdom over time. (e) Redline shows the number of deaths per day and the blue line shows how much ICU beds are required in peak days. (f) Redline shows the number of cases hospitalized per day and the blue line shows how much hospital beds are required in peak days.

### 4.6 SEIHCRD Model for the United States

The WHO health has reported that the medical facility of the USA is good and they have more hospital beds and ICU beds for people in comparison to other countries by ratio. United States has the highest number of COVID-19 cases in the world. According to the SEIHCRD model for the United States of America COVID-19, cases are on a peak between 130 and 145 days after that it is improving, its speed is very slow but steady and cases stop in October. New York, California, Illinois, and New Jersey have the most affected states of the USA. The proposed model for the USA has shown that between 120 and 180 days, the number of hospital beds and the number of ICU beds required but its count is very low. That time the number of death has 1200 per day and daily-hospitalized cases have nearly 1400. The result shows the basic reproduction rate is 4.5 in starting after some time it decreases and goes up to one and even lower. The case fatality rate goes up to 8.0, after 200 days it drastically fell. Our proposed method shows that the USA has the highest number of cases therefore, it takes more time in recovery. SEIHCRD Model results for the United States has shown in Fig. 23.

**Figure 23:**
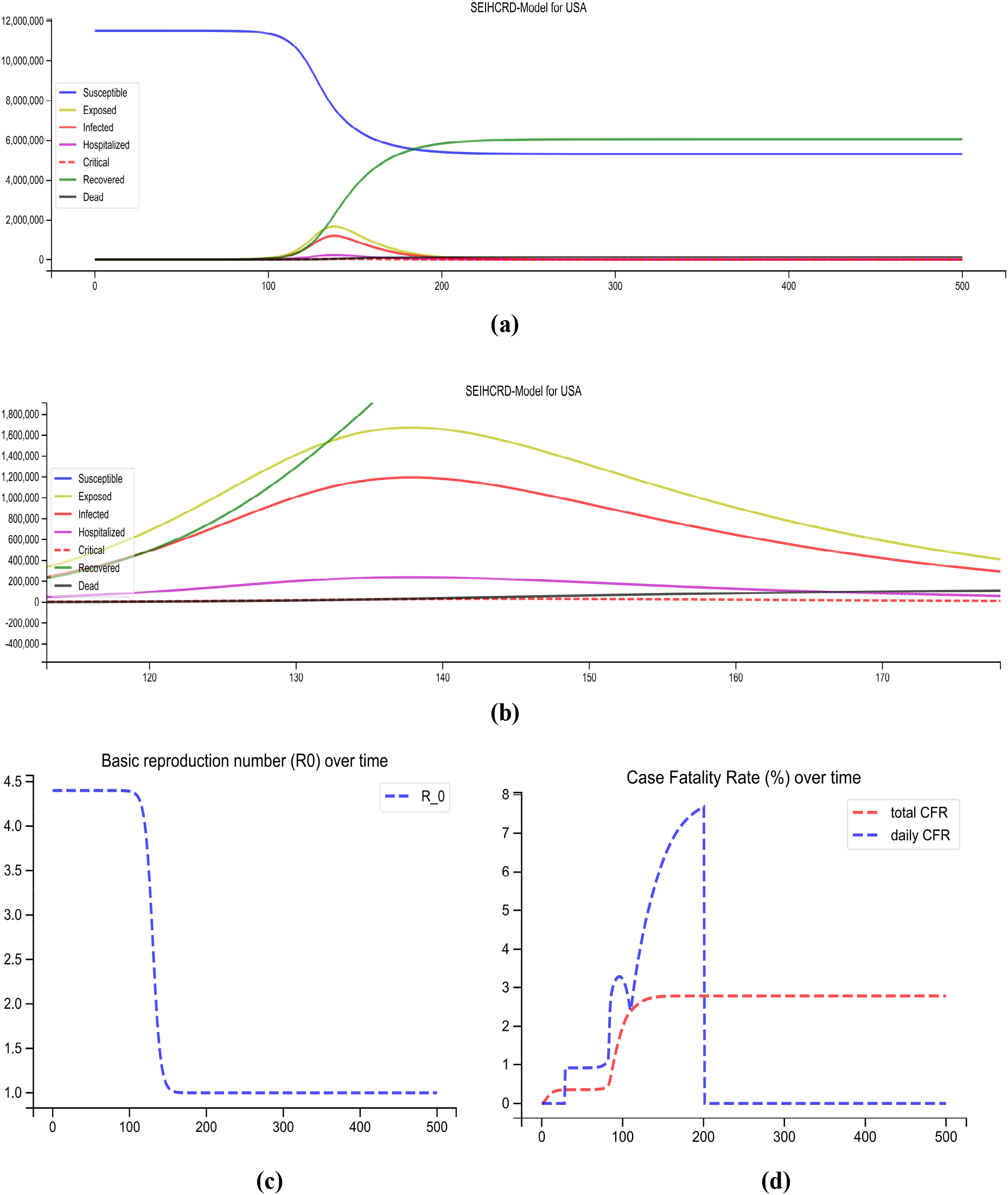
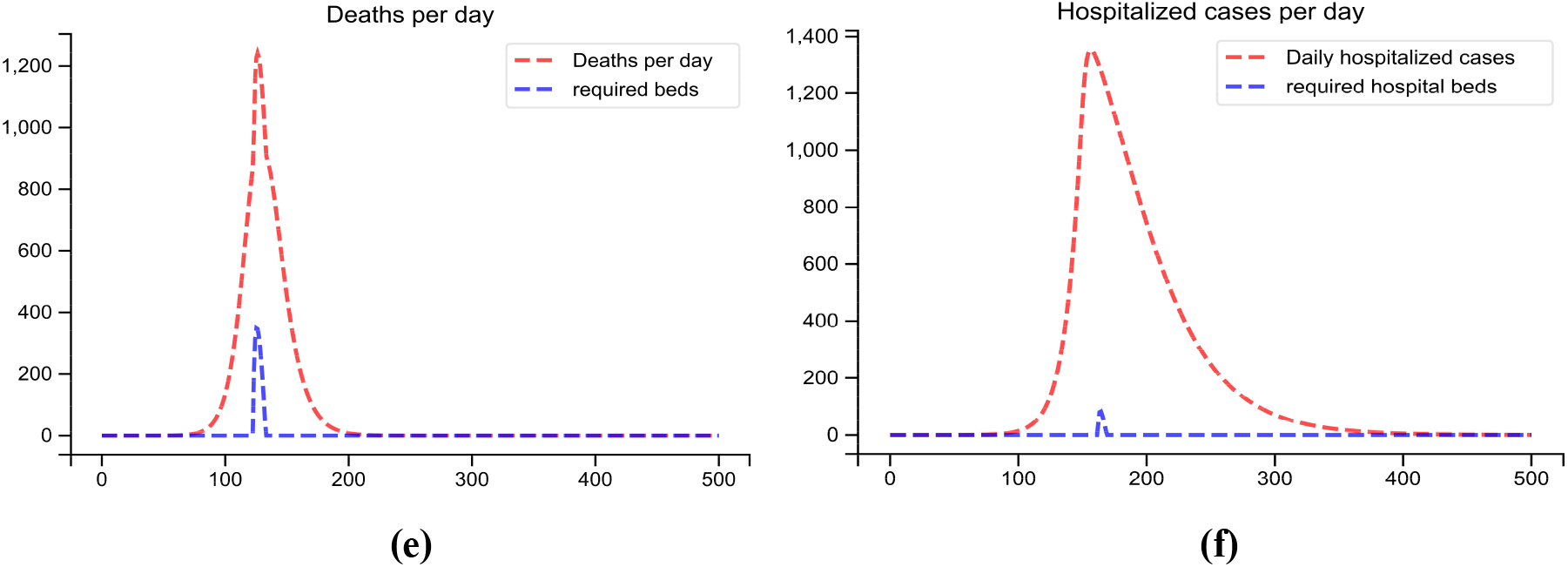
SEIHCRD Model for the United States (a) Spread scenario of SEIHCRD Model for the United States. A 500-day analysis has been done by the proposed model, which starts from 22 January. (b) In this, the peak point of the model has shown by zooming for cases. (c) The basic reproduction number of the United States over time (d) The case fatality rate of the United States over time. (e) Redline shows the number of deaths per day and the blue line shows how much ICU beds are required in peak days. (f) Redline shows the number of cases hospitalized per day and the blue line shows how much hospital beds are required in peak days.

## Limitations

Models are always simplifications of the real world. No models are perfect; there are some shortcomings in it. SEIHCRD Model also has some limitations, which are as follows:

Our system of differential equations is very sensitive to initial parameters. We have to very careful while given the initial parameters. Small changes in parameters can cause a huge difference in results. Our present manuscript has especially focused on serious cases and death cases. We have consented the serious cases that do not get treatment in the critical compartment. We have considered recovered cases not infected again in the future. R_0_ value cannot be increased; it either decreases or remain constant.

## Conclusion

The mathematical model is very helpful to predict the disease outbreak of COVID-19 and the SIR model and SEIR model have been widely used for prediction. Our proposed SEIHCRD model is an extension of the SEIR model in which three-compartments has added that is death, hospitalized, and critical. People with a serious infection, suffer from severe pneumonia need hospitalization. These individuals may either recover or progress to the critical compartment. People with critical infection experience multi-organ failure and multiple disorders require treatment in an ICU. These people either recover from the disease or die from it.

The SEIHCRD model estimates the date and magnitude of peaks of corresponding to exposed people, the number of infected people, the number of people hospitalized, the number of people admitted in ICUs, and the number of death of COVID-19. The SEIHCRD model is time-dependent hence; we have calculated the number of cases infected cases, hospitalized cases, and critical cases with time. We have data of hospital beds and ICU beds of most infected countries by COVID-19 after that we have calculated the hospitalized cases and critical cases using the proposed model then after calculation we can say how much beds are required. SEIHCRD-Model also computes the basic reproduction number over time, which is nearly the same as real data, but sometimes it varies, we also compute the case fatality rate over time of COVID-19 most affected countries. The model computes two types of Case fatality rate one is CFR daily and the second one is total CFR.

SEIHCRD model has shown, Spain and Italy have seen their worst times. The result shows that COVID-19 cases of Spain have ended completely in June last or July starting. Italy will be recovered completely by this disaster in August. India and Brazil have not yet seen the peak of the disaster. The result shows COVID-19 cases are at a peak in June in India and it is on a peak in June and July in Brazil. India needs some hospital beds in June. Brazil is going to have a shortage of hospital beds and ICU beds in June and July. The United State of America has suffered this tragedy the most. The USA has the highest number of total cases and deceased cases. The USA takes more days to recover and it will be recovered in October. The SEIHCRD model has shown the United Kingdom has also suffered this tragedy a lot, and it will be recovered in August.

## Data Availability

we give all information about data from where it takes.

## Acknowledgment

The work has been supported by a grant received from the Ministry of Education, Government of India under the Scheme for the Promotion of Academic and Research Collaboration (SPARC), 2019.

